# The responsiveness of health service provider and quality of services when provided at three selected urban primary health care sites in Dhaka city, Bangladesh: A cross–sectional study

**DOI:** 10.1101/2022.03.28.22273063

**Authors:** Abhiyan Gautam, Bandana Bhandari, Chungano Hassan, Fatiha Hassan, Saima Mehjabeen, Syed Masud Ahmed, Kuhel Faizul Islam, Nahitun Naher, Roksana Haque

## Abstract

1

**Introduction:** Responsiveness of Health Service Provider (HSP) and quality of services when provided resembles basic professional and social duties of HSP towards their clients. Because of poor responsiveness and quality of services when provided, clients lose their trust towards HSP. These factors are very important to improve relationship between HSP and clients, clients’ satisfaction, quality of care and finally increase utilization of Urban Primary Health Care Centre services (UPHC).

**Objectives:** This study was done to determine the responsiveness of health service provider and quality of services when provided at selected UPHCs in Dhaka city.

**Methodology:** A cross sectional quantitative study was conducted in three UPHCs in Dhaka city from November to December 2017. 257 exit interviews were conducted by systematic random sampling for responsiveness and quality of services when provided. 49 observations of client-provider interactions were conducted using Responsiveness of Physician (ROP) scale. For exit interview, dichotomous variable was used. Descriptive analysis was done using Stata v 12.1.

**Findings:** Majority (90%) of HSP listen carefully, explained about the diseases, facilitated about follow-up, and client understood information clearly. More than 70% of the clients found the providers approach were friendly though only 37% had social talk with the clients. 41% of the clients reported that the providers shared emergency contact number. Around 67% of clients were not asked allergic history and in 47% case consent was not taken before procedure. Being urban area, for more than 39% clients services were not given similar in terms of social status like gender, ethnicity, economic and social status.

For tangible items like gloves (80%) and thermometer (55%) were mostly missing in all UPHCs. 88% of the HSP were reliable, 93% assured the client and 91% showed empathy in all facilities. Clients were mostly satisfied with doctor’s behaviour and dissatisfied about the long waiting time (average 37 minutes) in all UPHCs.

**Conclusion:** This study has highlighted some important gaps in responsiveness of HSP which translate into the quality of care being provided to clients seeking care from UPHC. Friendliness of HSP should be increased and services should be provided with respect.

## 2. Introduction

Responsiveness of HSP and quality of services when provided resembles basic ethical and social duties of HSP towards their clients. It has inter relation with WHO health system building [1, 2, 3].

Responsiveness is related with protection and continuation of clients’ rights to quality, adequate, and timely care [4]. It is a combination of human rights, medical ethics, and human development [5].

Poor responsiveness can discourage clients to take interest on health related issues and early consultation. Lack of information, confidentiality, privacy, female attendants for female clients, friendly behavior, safety, equitable services, delay in treatment, trust, dignity, respect, clear communication, and high expectation, service charge etc. from can influence perception towards HSP [6, 7]. On other hand, high patient flow, health system limitation, lack of communication skills of providers and disrespect by patient influence responsiveness from HSP side [8]. Health services resources such as high coverage area, poor governance, coordination and information sharing mechanism, lack of infrastructure, equipment, drugs, and less staffs were the major responsible factors which indirectly influence responsiveness. Loss of trust between providers and clients can leads to low utilization of services [9]. Every unit were responsible for responsiveness and quality of services when provided.

Quality of services when provided refers to affordable, effective, safe and standard services by skilled HSP which satisfy clients [10]. Technical competence, client care, management, environment safety, tangibles, reliable, responsiveness, assurance, empathy, and clients’ satisfactions influence quality of services [10, 11, 12]. Reliability of HSP has the highest value where tangibles has the lowest values [6]. Lack of these integrated package influences on utilization of services.

LGD has mentioned that there is low finance allocation for health. With compromised budget in UPHC, manpower, equipment, and services can be compromised which in turn, will affect service delivery. Lack of regular refresher training and supervision from higher authority has raised questions on quality of services. Since there is no regular coordination between MoHFW and MoLGRDC, there is gap in technical advice to improve quality of services when provided [13, 14, 15].

In Bangladesh, conflicts between doctors and clients is increasing because patient claims that doctors neglect patient while doctors claim back that they are assaulted by clients [8]. Factors like behaviour, friendliness, respecting, informing and guiding, gaining trust and optimizing benefit etc. which influence responsiveness and this directly affect the quality of services when provided at health facilities [7, 16, 17, 18, 19, 20, 21].

### Justification

Despite establishment of UPHC services mechanism and having national urban health strategy and guidelines health indicators of urban population remain still poor in the country. Due to lack of coordination between MoHFW, NGOs and MoLGRDC; the monitoring and evaluation of the services remain constrained [13, 22, 23]. HSP are key persons to guide and satisfy their clients, and to improve quality of care. Though clients visiting health facilities have high expectations, but level of satisfaction is found to be low [6, 24]. There are limited studies on stated aspects. Since responsiveness and quality of services both influence the service access and utilization, hence exploring responsiveness and quality is crucial to ensure service use and promote country achieve sustainable development goals. Therefore, this study will help to give evidence to improve policy and fill the knowledge gaps.

### Conceptual Framework

In UPHCs, organizational structure, facilities and provider’s perspectives can influence on responsiveness of HSP towards clients and quality of services when provided at UPHC. Responsiveness is determined by information, friendliness, trustworthiness, non-maleficence and respect for dignity of person. Similarly, quality of services when provided is determined by tangible items, reliability, assurance, empathy, and client satisfaction. These factors jointly influence on utilization of the services (Fig 1).

**Figure 1:**
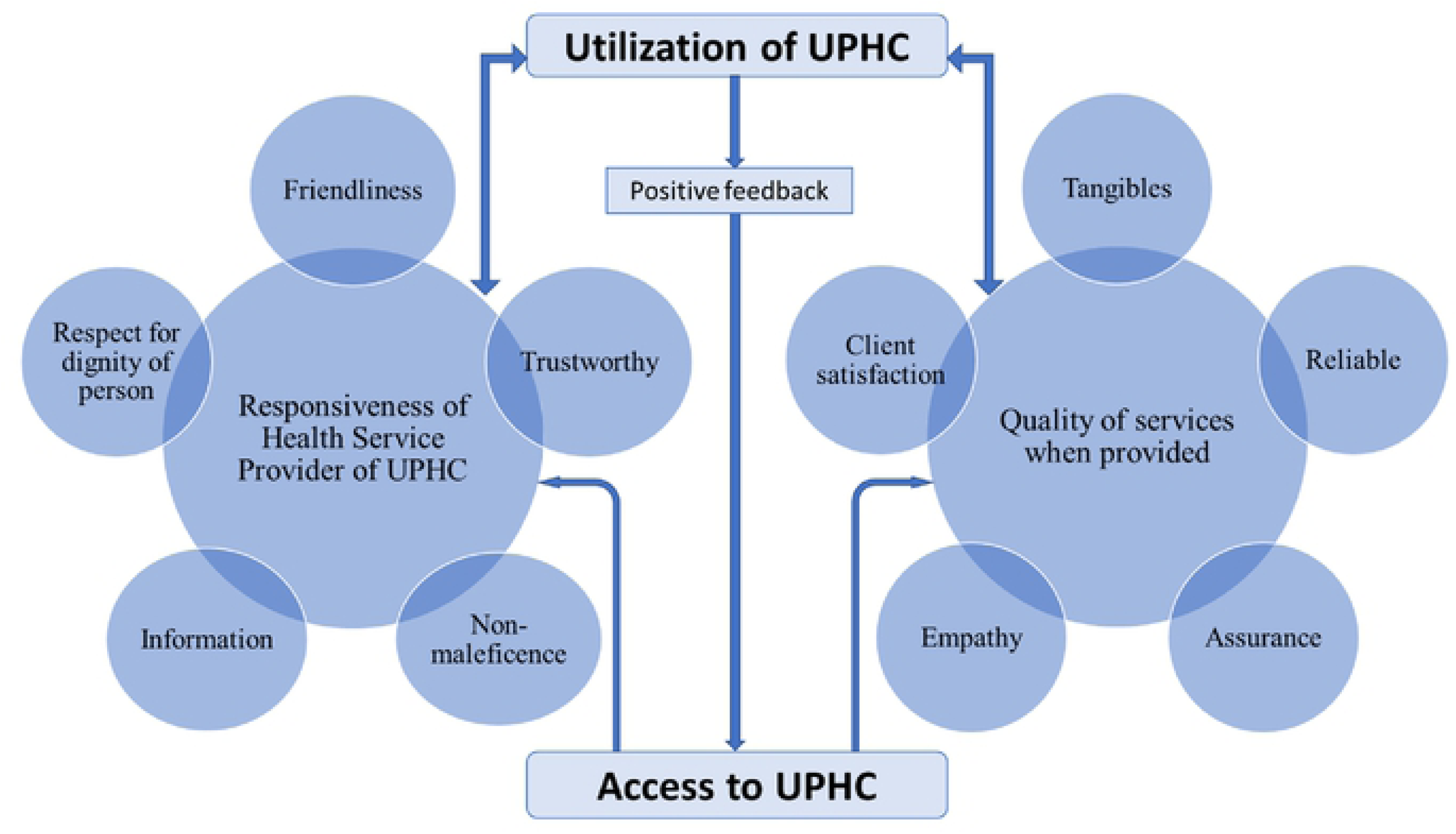
Conceptual Framework

### Objective

#### Specific Objectives

a. To measure responsiveness of health service provider towards clients in UPHC.
b. To find level of quality of services when provided by HSP in UPHC.

## 3. Methodology

### Study Design and Approach

The study had cross-sectional design with a quantitative approach. Quantitative approach was preferred to collect detailed description on responsiveness of HSP and quality of care when provided at UPHC.

### Study Site

We reviewed list of partner NGOs of MoLGRDC, UPHC project. There are 25 NGOs who are providing UPHC services on behalf of LGB. Among them, 10 organizations are providing UPHC services within Dhaka City Corporation areas. Out of them, three partners NGOs are selected. KMSS and Nari Maitree from DNCC and BAPSA from DSCC. We selected them according to our convenience. UPHCs were selected both from Dhaka North and South City Corporation to have rich picture of Dhaka.

These UPHCs provide low cost primary health care services to 3 to 4 lakhs of urban people. Their services are mainly focused to poor and vulnerable population of slum area. It gives more subsidies to poor people in the form of red card.

UPHC has three types of services delivery centers; CRHCC, PHCC and satellite clinic. Catchment area of each UPHC ranges from 4 to 7 wards. One UPHC has a CRHCC which mainly focuses on maternal and child health services. Under a CRHCC there are 4 to 7 PHCCs one in each ward. There are around 80 satellite clinic pockets in each UPHC. It provides services from 15 sites in each day (Fig 2).

**Figure 2.**
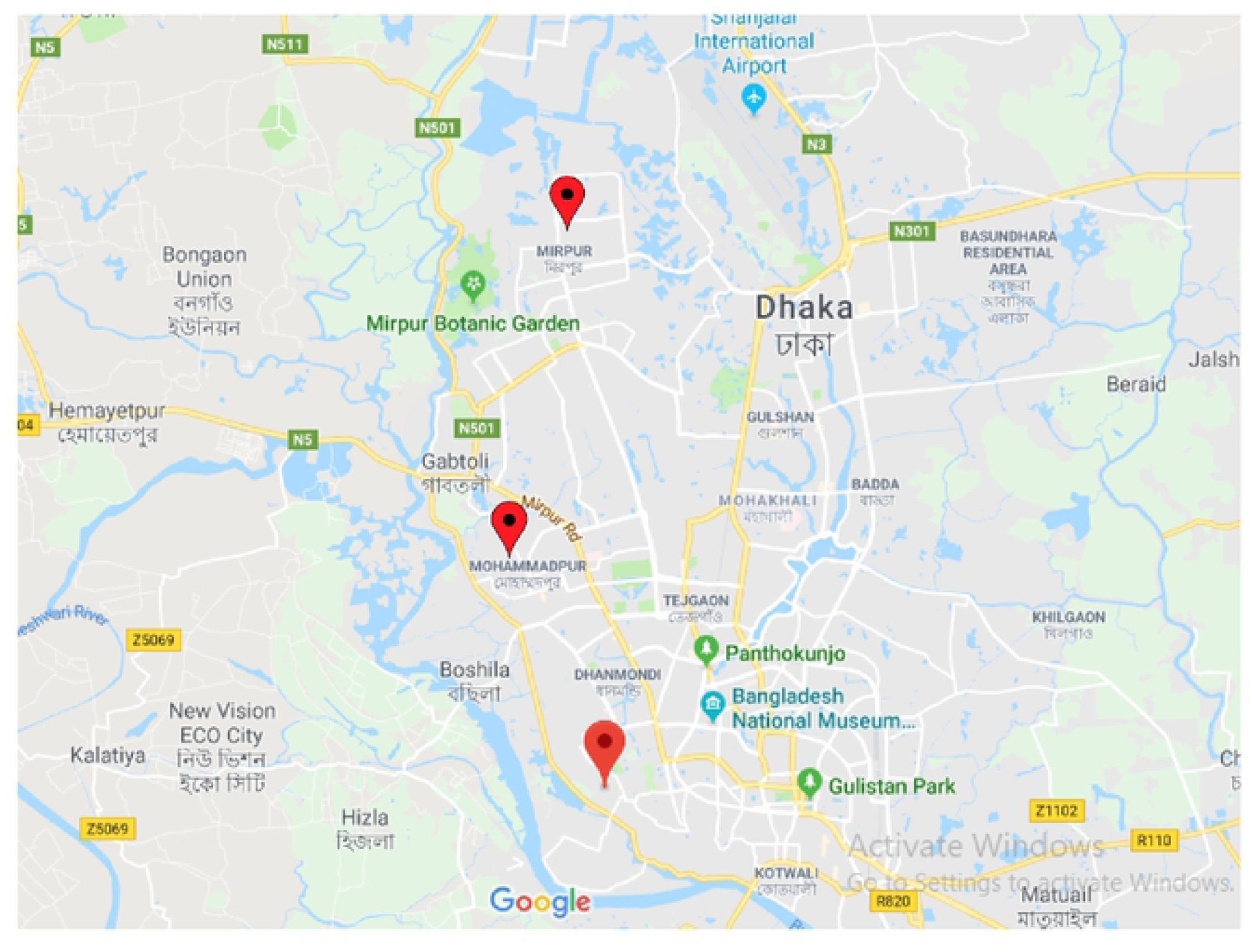
Map showing study sites (order from top to bottom; KMSS, Nari Maitree and BAPSA) Study sites I. Dhaka South City Corporation:

i. Bangladesh Association for Prevention of Septic Abortion (BAPSA), Hazaribag park, Hazaribag, Dhaka II. Dhaka North City Corporation:

i. Nari Maitree, Baitul Aman Cooperative Housing Society Ltd, Ring Road, Dhaka
ii. Khulna Mukti Sewa Sangstha, Mirpur, Dhaka

### Study participants

We selected service users for exit interview and service providers for observation as study participants in CRHCC and PHCC of the selected UPHCs.

Study participants were service users of selected UPHCs. For client-provider interaction, service providers were observed.

### Inclusion and Exclusion Criteria

Clients, 18 years and above utilizing UPHC of the selected centres were included in the study. For below 18 years clients, consent were taken with their parents. Intellectually challenged and clients in critical condition were excluded from the study.

### Sampling Technique and Sample Size

In this research, systematic random sampling technique was used where sampling unit was clients of UPHC. Study was done in three different UPHCs. Every 4^th^ outdoor patient of clients UPHC was selected randomly for the exit interview. Sample size of our study was 384 by using proportionate sampling methods but we were able to collect only 257. We did 49 observation of 7 HSP.

Proportionate Sample Size Formula: n=z^2^ * v (1-v)/ d^2^ where v= p_1_ (1-p_1_) + p_2_ (1-p_2_)

Here, Utilization rate of Nari Maitree, DNCC (P_1_) = 54%

Utilization rate of BAPSA, DSCC P_2_ = 44%

By keeping Z= 1.96 and level of error (d) =0.05,

Hence, Sample size (N) = 384

### Study Tools

#### Tool development

The study was done by using structured questionnaire and observation checklist (ROP scale) for responsiveness and quality of services when provided. The tools were adopted from a study done by Joarder et al. in 2017 using ROP scale for responsiveness component [20]. We developed preliminary set of questionnaire in English. We did validation followed by multiple discussion with supervisors and expert in the same field. Since ROP scale tool was validated for rural part, our tool was modified according to the context of Dhaka. Elements of ROP scale was kept as a variable for exit interview. Score of ROP scale (0 to 4) was modified to dichotomous scale (Yes or No). In place of financial sensitivity, we added non-xmaleficence. But for observation, same ROP-scale tool was used which was developed by Joarder et al. It was done for content validity.

Similarly, we also adapted SERVQAL tool for quality of services when provided [25, 26]. It was also modified to dichotomous scale (Yes or No). We used same domains and variables from the adopted tool, we added client satisfaction and dissatisfaction domains to know what kind of services clients get satisfy from.

#### Pre-testing and finalizing tool

Preliminary questionnaire was pre-tested to ensure intelligibility, length, organization and structure of proposed questionnaire to the clients of Nari Maitree. After getting feedback of pre tested questionnaire, we again modified to give fine-tune in the questions of various domains of two objectives. To make same level of understanding among the researchers and research assistants, we made Bengali version of the same questionnaire. However, we collected data on English version.

### Data collection

We collected data by interviewing service users and observing when service was provided by using the tools from 19 November to 3 December 2017.

Structured questionnaire was used during exit interview to collect clients’ experiences on responsiveness and quality of services provided by HSP because it will give immediate feedback after utilizing the services. The tool was interviewer assisted. Side by side, observation was done to triangulate the data. ROP scale was used to observe the responsiveness. Five variables for each responsiveness of HSP and quality of services when provided have been selected. (Annex 1 & 2)

For exit interview, Bengali speaking research assistant was assisting because researcher was non-Bengali speaking international student.

### Data Entry and Analysis

#### Ensuring data quality

We explained to our research assistants and researchers about our questionnaire in detail. We translated our tool in Bangla version and in each question, we gave examples so that all researchers’ have same level of understanding. We also checked data collection methods and integrity by searching missing values (completeness) in data. We have followed systematic random sampling method to collect data to reduce selection bias.

#### Data entry and analysis

Data were entered in google form and imported into MS excel where data cleaning was done. Missing values were imputed by using multiple imputation in Stata version 12.1. Univariate analysis was also done.

Data of three UPHCs were separated. Again, domains of each specific objectives were segregated. Variables under each domain were put together to make it easy to calculate total scores. In exit interview, we asked service users regarding their perception regarding HSP. It was dichotomous variable with yes or no answer. (Table 2 & 3) Positive and negative answers were calculated separately. We used descriptive analysis method [12, 26].

**Table 2.**
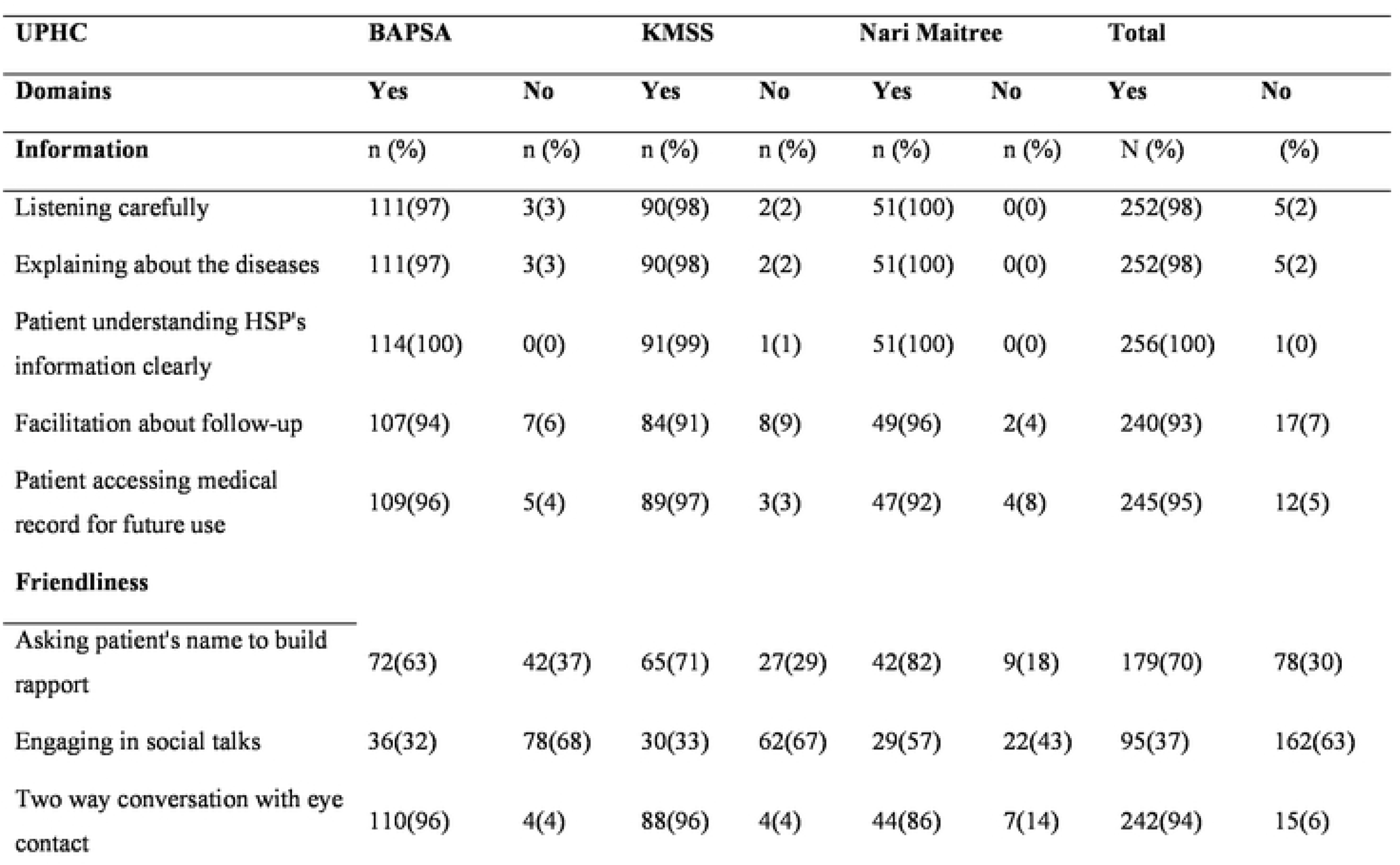
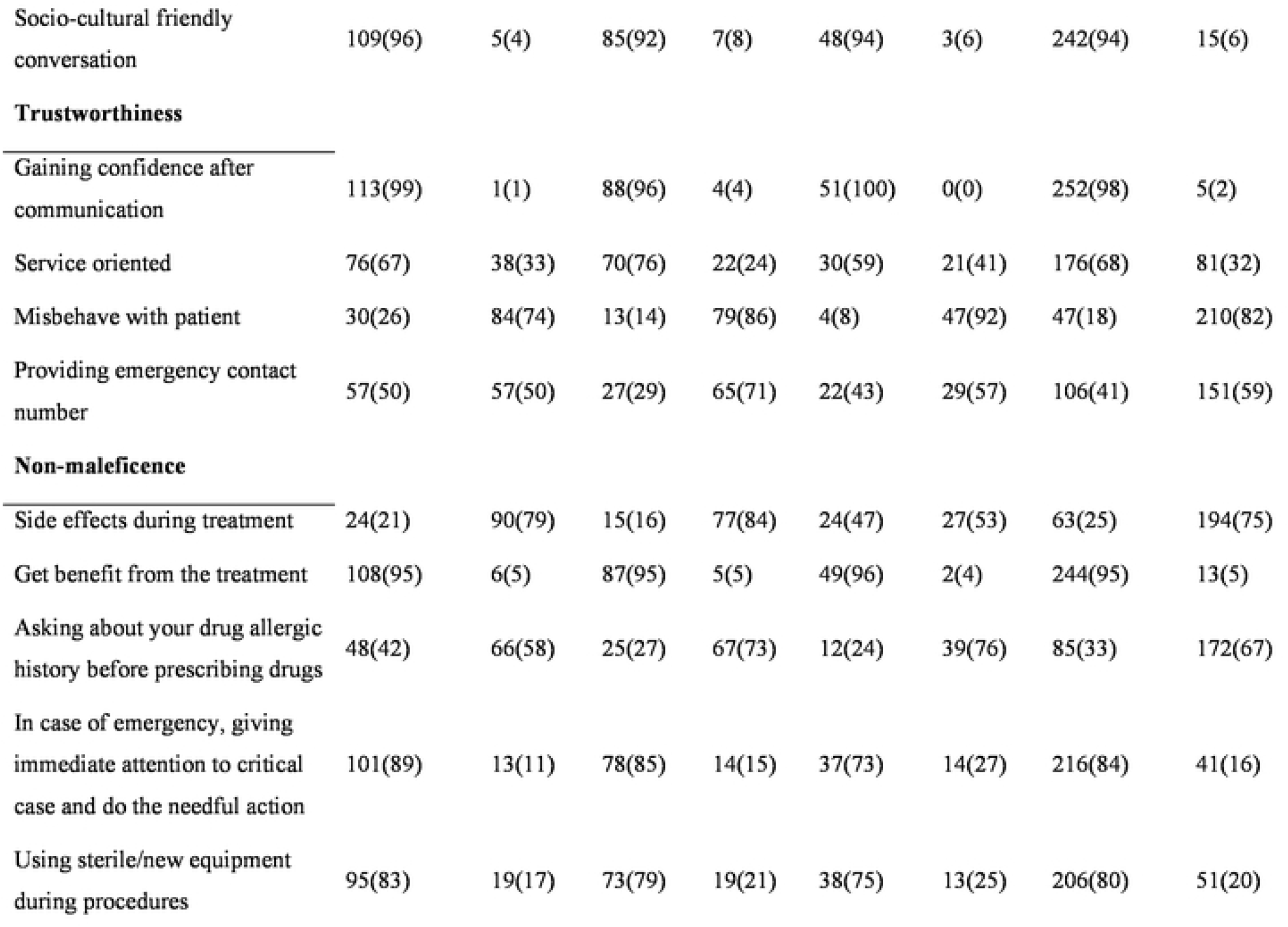
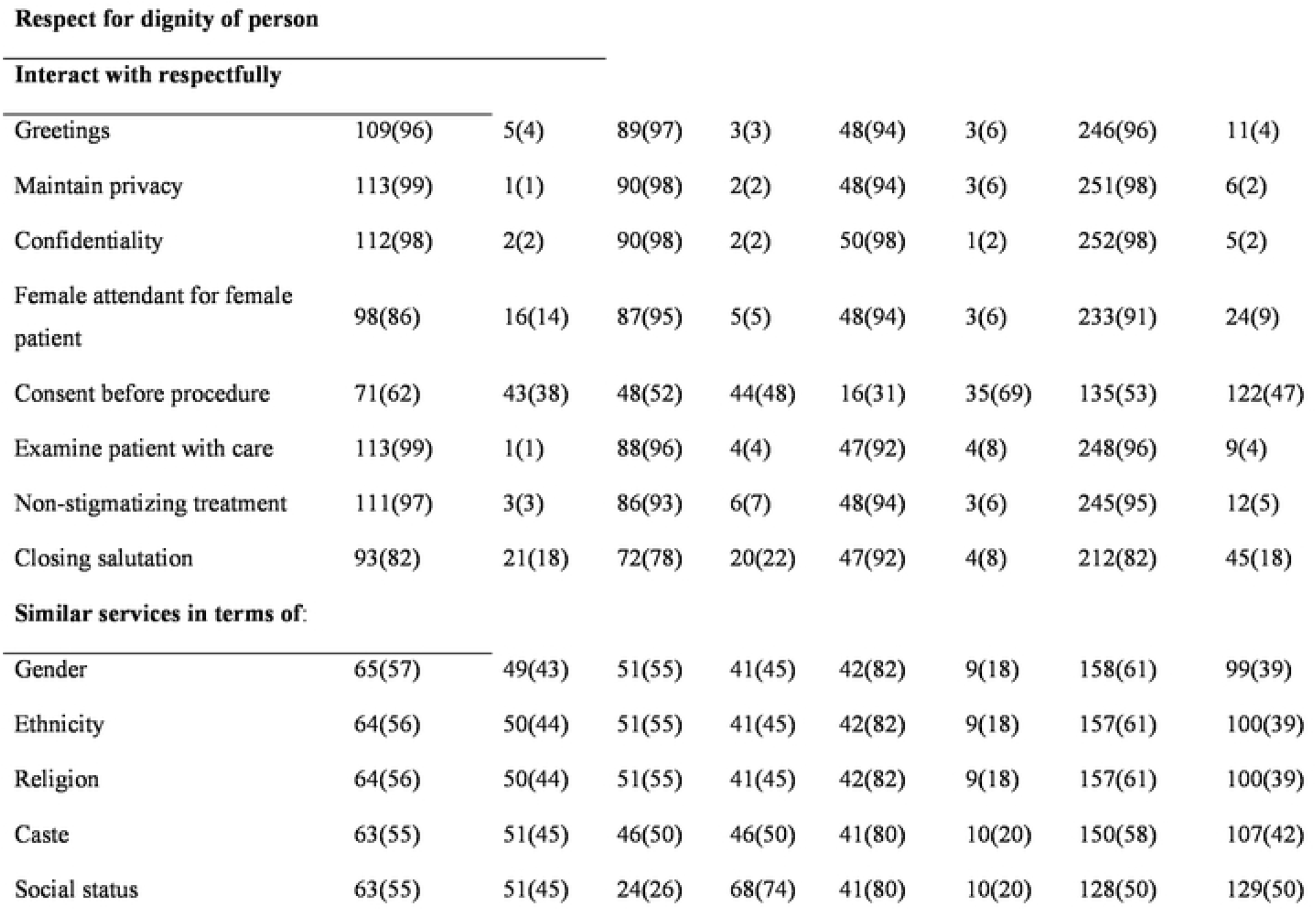
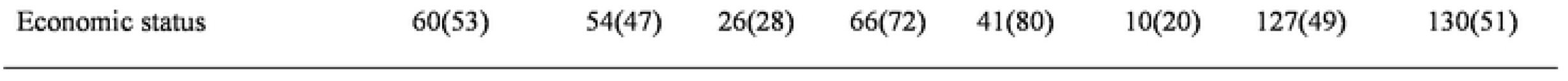
Responsiveness of Health Service Provider (HSP) in three UPHCs

| UPHC | BAPSA |  | KMSS |  | Nari Maitree |  | Total |  |
| --- | --- | --- | --- | --- | --- | --- | --- | --- |
| Domains | Yes | No | Yes | No | Yes | No | Yes | No |
| Information | n (%) | n (%) | n (%) | n (%) | n (%) | n (%) | N (%) | (%) |
| Listening carefully | 111(97) | 3(3) | 90(98) | 2(2) | 51(100) | 0(0) | 252(98) | 5(2) |
| Explaining about the diseases | 111(97) | 3(3) | 90(98) | 2(2) | 51(100) | 0(0) | 252(98) | 5(2) |
| Patient understanding HSP's information clearly | 114(100) | 0(0) | 91(99) | 1(1) | 51(100) | 0(0) | 256(100) | 1(0) |
| Facilitation about follow-up | 107(94) | 7(6) | 84(91) | 8(9) | 49(96) | 2(4) | 240(93) | 17(7) |
| Patient accessing medical record for future use | 109(96) | 5(4) | 89(97) | 3(3) | 47(92) | 4(8) | 245(95) | 12(5) |
| **Friendliness** |  |  |  |  |  |  |  |  |
| Asking patient's name to build rapport | 72(63) | 42(37) | 65(71) | 27(29) | 42(82) | 9(18) | 179(70) | 78(30) |
| Engaging in social talks | 36(32) | 78(68) | 30(33) | 62(67) | 29(57) | 22(43) | 95(37) | 162(63) |
| Two way conversation with eye contact | 110(96) | 4(4) | 88(96) | 4(4) | 44(86) | 7(14) | 242(94) | 15(6) |
| Socio-cultural friendly conversation | 109(96) | 5(4) | 85(92) | 7(8) | 48(94) | 3(6) | 242(94) | 15(6) |
| **Trustworthiness** |  |  |  |  |  |  |  |  |
| Gaining confidence after communication | 113(99) | 1(1) | 88(96) | 4(4) | 51(100) | 0(0) | 252(98) | 5(2) |
| Service oriented | 76(67) | 38(33) | 70(76) | 22(24) | 30(59) | 21(41) | 176(68) | 81(32) |
| Misbehave with patient | 30(26) | 84(74) | 13(14) | 79(86) | 4(8) | 47(92) | 47(18) | 210(82) |
| Providing emergency contact number | 57(50) | 57(50) | 27(29) | 65(71) | 22(43) | 29(57) | 106(41) | 151(59) |
| **Non-maleficence** |  |  |  |  |  |  |  |  |
| Side effects during treatment | 24(21) | 90(79) | 15(16) | 77(84) | 24(47) | 27(53) | 63(25) | 194(75) |
| Get benefit from the treatment | 108(95) | 6(5) | 87(95) | 5(5) | 49(96) | 2(4) | 244(95) | 13(5) |
| Asking about your drug allergic history before prescribing drugs | 48(42) | 66(58) | 25(27) | 67(73) | 12(24) | 39(76) | 85(33) | 172(67) |
| In case of emergency, giving immediate attention to critical case and do the needful action | 101(89) | 13(11) | 78(85) | 14(15) | 37(73) | 14(27) | 216(84) | 41(16) |
| Using sterile/new equipment during procedures | 95(83) | 19(17) | 73(79) | 19(21) | 38(75) | 13(25) | 206(80) | 51(20) |
| **Respect for dignity of person****Interact with respectfully** |  |  |  |  |  |  |  |  |
| Greetings | 109(96) | 5(4) | 89(97) | 3(3) | 48(94) | 3(6) | 246(96) | 11(4) |
| Maintain privacy | 113(99) | 1(1) | 90(98) | 2(2) | 48(94) | 3(6) | 251(98) | 6(2) |
| Confidentiality | 112(98) | 2(2) | 90(98) | 2(2) | 50(98) | 1(2) | 252(98) | 5(2) |
| Female attendant for female patient | 98(86) | 16(14) | 87(95) | 5(5) | 48(94) | 3(6) | 233(91) | 24(9) |
| Consent before procedure | 71(62) | 43(38) | 48(52) | 44(48) | 16(31) | 35(69) | 135(53) | 122(47) |
| Examine patient with care | 113(99) | 1(1) | 88(96) | 4(4) | 47(92) | 4(8) | 248(96) | 9(4) |
| Non-stigmatizing treatment | 111(97) | 3(3) | 86(93) | 6(7) | 48(94) | 3(6) | 245(95) | 12(5) |
| Closing salutation | 93(82) | 21(18) | 72(78) | 20(22) | 47(92) | 4(8) | 212(82) | 45(18) |
| **Similar services in terms of:** |  |  |  |  |  |  |  |  |
| Gender | 65(57) | 49(43) | 51(55) | 41(45) | 42(82) | 9(18) | 158(61) | 99(39) |
| Ethnicity | 64(56) | 50(44) | 51(55) | 41(45) | 42(82) | 9(18) | 157(61) | 100(39) |
| Religion | 64(56) | 50(44) | 51(55) | 41(45) | 42(82) | 9(18) | 157(61) | 100(39) |
| Caste | 63(55) | 51(45) | 46(50) | 46(50) | 41(80) | 10(20) | 150(58) | 107(42) |
| Social status | 63(55) | 51(45) | 24(26) | 68(74) | 41(80) | 10(20) | 128(50) | 129(50) |
| Economic status | 60(53) | 54(47) | 26(28) | 66(72) | 41(80) | 10(20) | 127(49) | 130(51) |

**Table 3.** Average responsiveness scores according to ROP scale

| Domains | Variables | Average Scores<sup>a</sup> |
| --- | --- | --- |
| Friendliness | Ask Patient's Name | 1.9 |
|  | Social Talks | 2.0 |
|  | Ask about family | 1.5 |
|  | Friendly | 2.0 |
|  | Use of patient friendly language | 2.7 |
|  | Sense of Humor | 1.7 |
| Information | Write after listening | 2.4 |
|  | Listen to patient | 2.7 |
|  | Suggestion for Prevention | 2.2 |
|  | Follow up advice | 2.0 |
|  | Explain about diseases | 2.2 |
|  | Patient understand suggestion | 1.8 |
| Respect | Greet and welcome to patient | 2.1 |
|  | Physical examination | 2.1 |
|  | Behavior of HSP | 2.2 |
|  | Saying good-bye | 1.8 |
| Trustworthiness | Misbehave | 2.6 |
|  | Service oriented | 3.2 |
|  | Illegal activities of HSP | 3.9 |

<sup>a</sup>Out of total score of 4

Frequencies and percentages were calculated and differences among the UPHCs were calculated. For some variables (e.g. waiting time and consultation time) of quality of services, range was identified and plotted in graphs to see the differences among the three UPHCs. For variables like satisfaction and dissatisfaction, we merge similar answer into three subtopic and plotted graphs and compare among three facilities.

Similarly, during observation we score from 0 to 4 according to the example we had in observation checklist, then we find mean score of each variable under respective domains of responsiveness. Then mean value was calculated.

### Ethical consideration

Ethical authorization was obtained from the Institutional Review Board (IRB), JPGSPH on 15^th^ November 2017. As an enlisted organization to provide UPHC services, prior approval from MoLGRDC was obtained to collect data from the selected UPHC centres. For every participant, informed consent was taken. Purpose of study, risk and benefit was explained before proceeding for the interviews. Participants could withdraw questions and skip the study at any time during the study if they wanted. We assured that participants name would not be revealed, privacy and confidentiality would maintain. Then we took written informed consent before interview. Similarly, for observation, we took consent of HSP. (Annex 3)

## 4. Findings

### Sociodemographic of respondents

Our research was aimed to find the quality of services when provided and responsiveness of HSP by using exit interview. Out of 257 exit interviews, 114 were from BAPSA, 92 from KMSS and 51 were from Nari Maitree. In our study, 98.8% respondents were female and 1.2% were Male. Around 5% participants were below 18 years, more than 50% were from 18 to 24 age group, 33% were from 25 to 34, rest of them were above 35 years. About 98% were married. In total, 95% follow Islam religion. Only, 10% were employed. Around 82% clients came from catchment area of respective UPHCs. Only 7.3% were uneducated. We did descriptive analysis. Our findings are divided into two main parts according to the objectives.

### Findings related to responsiveness of HSP and quality of care when provided

The study findings are presented as per variables used under each domains of responsiveness of HSP and quality of service when provided. (Table 1)

**Table 1.** Variables under each Domains

| **Responsiveness of health service provider** | **Quality of services when provided** |
| --- | --- |
| a. Information | a. Tangibles |
| b. Friendliness | b. Reliable |
| c. Trustworthiness | c. Assurance |
| d. Non-maleficence | d. Empathy |
| e. Respect for dignity of person | e. Client Satisfaction |

Note:
For responsiveness medical ethics, human rights and human development components were modified and adopted from “Health and Human Rights Working Paper, 2003” and “Medial Ethics Manual, World Medical Association, 2015”.
For quality, domains were modified and adopted from “SERVQUAL factors” and “service availability and readiness assessment (SARA)” developed by Zeithaml, Parasuraman and Berry, and WHO respectively (Zeithaml, 2009; WHO, 2013; Nallusamy, Hariharan & Vijaykumar, 2017).

### Responsiveness of HSP

ROP scale findings of exit interview and observations under five domains have been explained below simultaneously. (Table 2 and 3)

#### Information

On this domain, 98% clients in exit interview said that HSP listen them carefully and explain about the diseases. It was highest in Nari Maitree where 100% clients said HSP listen them carefully. From the observation, the average score on listen to patient was 2.7 out of 4 and score for HSP writing after listening to patient was 2.4 while explain about diseases was 2.2. HSP also suggested patient regarding preventive measures and its average score from ROP scale was 2.2. These observation score means HSP showed attentiveness to listen patient queries but after some stage, HSP stopped the patient and gone into the next step. HSP explained at least one of the things among name of diseases or treatment. Observation also showed that, HSP indirectly explained about preventive and health promotive measures.

100% of clients understood information clearly in all UPHCs. In contrast to that, our observation showed average score of clients understanding suggestion was only 1.8 out of 4. Here, in many cases HSP did not ask the patient about his understanding after explaining the information. For 7% of clients, facilitation was not given for follow up. From observation average ROP-scale score was 2 out of 4. That means, HSP gave minimum follow-up plans, date when to come again or what would be the cost of follow-up etc.

For 5%, patients were not able to access medical record for future use. From our observation, they HSP only wrote prescription in 100% cases but findings from their examination was not written in all cases. However, for ANC cases, some examination findings were written.

#### Friendliness

Only 70% clients said HSP asked their name and only 37% engaged in social talks to build rapport. In BAPSA, 37% of clients were not asked their name and for 68% social talks was not done as a part of rapport building. According to observation, the average score for asking patient’s name was 1.9, social talks was 2 out of 4. It means that, HSP asked the name but did not care about it or used it. There was a minimum social talk.

In few cases, HSP asked about the family during the talks. Its score was 1.5 out of 4. That means, HSP was not interested to know about family during their conversation.

Around 94% clients said that conversation was socio-cultural friendly and two-way conversation with eye contact in all facilities. Observation showed that average score of friendly was 2, use of patient friendly language was 2.7 and sense of humor was 1.7 out of 4. Friendliness scores showed that, HSP were friendly in minimum level. HSP used some medical terminologies and explained some of it. It also showed that, HSP had some humor or smiling during the conversation.

#### Trustworthiness

Nearly 98% patient gain confidence after communication and 41% clients got an emergency contact number of HSP at the end of communication in all UPHCs. In KMSS, 71% clients said that, emergency number was not given. Although, our observation showed that, emergency contact number was not given in any of the facilities. However, 32% clients found that HSP were not service oriented. Observation showed, average score of service orientation service was 3.2 out of 4. Observation showed that HSP services were not business oriented but was not even service oriented also.

In 18% cases HSP misbehaved with patient during communication. Height misbehave from HSP was perceived in BAPSA (26%). Observation score was 2.6 out of 4. It suggested that, HSP showed minimum respect to the patient.

Observation average score in HSP involving in ethical and legal activities was 3.9 out of 4. It showed HSP was not involve in any illegal activities, which increase trust among the clients.

#### Non-maleficence

Patient found that 84% were given immediate attention during emergency. Only for 33% clients, drug allergic history was asked before prescribing medicine. For 20% client sterile or new equipment was not used during procedure. After the treatment, 95% got benefit from the treatment but 25% get the side effect. In Nari Maitree 47% clients perceived side effect after the treatment. From our observation, in all UPHCs, drug allergic history was not asked.

#### Respect for dignity of person

During interaction, in 96% cases HSP greeted or replied greetings. Average score from observation was 2.1 out of 4. It suggested that, HSP expressed slight greeting before the conversation.

For 98%, privacy and confidentiality were maintained and for 91% female attendant was present for the female patient. However, in 47% cases, consent was not taken before procedure. Average score from observation was 2.1 out of 4. Score suggested that HSP examined patient at least for once.

96% clients said that examination was done with care. Similarly, for 95% non-stigmatizing treatment was given. ROP scale score for behavior of HSP was 2.2 out of 4. Observation suggested that, HSP slightly discouraged clients to ask question.

After the treatment in 82% cases, closing salute was given. ROP scale score was 1.8 out of 4 on closing salute. It means HSP did not greeted but replied back by gesture.

Services were not similar in terms of gender, ethnicity, religion, caste, social status and economic status. In terms of gender, ethnicity and religion, 39% felt services was not similar.

Similarly, for social status and economic status, more than 50% felt services were not similar. Around 42% patient felt dissimilar services in terms of caste.

### Quality of services when provided

In exit interviews, we asked quality of services to service users (clients) regarding their perception when service was provided. They were dichotomous variables with yes or no answer. Findings on five domains are described below. (Table 4)

**Table 4.** Quality of services when provided in three UPHCs

| UPHCs | BAPSA |  | KMSS |  | Nari Maitree |  | Total |  |
| --- | --- | --- | --- | --- | --- | --- | --- | --- |
| Domains | Yes | No | Yes | No | Yes | No | Yes | No |
| Tangibility | n (%) | n (%) | n (%) | n (%) | n (%) | n (%) | N (%) | N (%) |
| Blood Pressure Measuring instrument | 96(84) | 18(16) | 83(90) | 9(10) | 42(82) | 9(18) | 221(86) | 36(14) |
| Gloves | 31(27) | 83(73) | 12(13) | 80(87) | 9(18) | 42(82) | 52(20) | 205(80) |
| Thermometer | 57(50) | 57(50) | 36(39) | 56(61) | 22(43) | 29(57) | 115(45) | 142(55) |
| Stethoscope | 88(77) | 26(23) | 75(82) | 15(16) | 40(78) | 11(22) | 203(79) | 52(20) |
| Measurement tape | 48(42) | 66(58) | 34(37) | 58(63) | 25(49) | 26(51) | 107(42) | 150(58) |
| Replacement of items after use | 64(56) | 50(44) | 37(40) | 55(60) | 26(51) | 25(49) | 127(49) | 130(51) |
| Doctors room visually attractive | 111(97) | 3(3) | 86(93) | 6(7) | 48(94) | 3(6) | 245(95) | 12(5) |
| **Reliability** |  |  |  |  |  |  |  |  |
| Responded timely | 107(94) | 7(6) | 89(97) | 3(3) | 51(100) | 0(0) | 247(96) | 10(4) |
| Writing on OPD records | 103(90) | 11(10) | 72(78) | 20(22) | 50(98) | 1(2) | 225(88) | 32(12) |
| Dependable | 113(99) | 1(1) | 88(96) | 4(4) | 50(98) | 1(2) | 251(98) | 6(2) |
| **Assurance** |  |  |  |  |  |  |  |  |
| Give reassurance | 110(96) | 4(4) | 87(95) | 5(5) | 51(100) | 0(0) | 248(96) | 9(4) |
| Safe to share problems | 112(98) | 2(2) | 88(96) | 4(4) | 50(98) | 1(2) | 250(97) | 7(3) |
| Assistance to help HSP | 104(91) | 10(9) | 87(95) | 5(5) | 47(92) | 4(8) | 238(93) | 19(7) |
| **Empathy** |  |  |  |  |  |  |  |  |
| Provide dedicated attention during consultation | 112(98) | 2(2) | 90(98) | 2(2) | 51(100) | 0(0) | 253(98) | 4(2) |
| Consultation time |  |  |  |  |  |  |  |  |
| Enough time to share problems | 104(91) | 10(9) | 82(89) | 10(11) | 47(92) | 4(8) | 233(91) | 24(9) |
| Able to identify health needs | 109(96) | 5(4) | 82(89) | 10(11) | 47(92) | 4(8) | 238(93) | 19(7) |
| Compassionate | 113(99) | 1(1) | 87(95) | 5(5) | 48(94) | 3(6) | 248(96) | 9(4) |
| **Client satisfaction** |  |  |  |  |  |  |  |  |
| Find all service | 47(41) | 67(59) | 30(33) | 62(67) | 22(43) | 29(57) | 99(39) | 158(61) |
| Availability of medicine | 99(87) | 15(13) | 67(73) | 25(27) | 41(80) | 10(20) | 207(81) | 50(19) |
| Availability of doctor | 83(73) | 31(27) | 65(71) | 27(29) | 44(86) | 7(14) | 192(75) | 65(25) |
| Availability of diagnostic test | 88(77) | 26(23) | 79(86) | 13(14) | 43(84) | 8(16) | 210(82) | 47(18) |
| Availability of services | 49(43) | 65(57) | 23(25) | 66(72) | 24(47) | 25(49) | 96(37) | 156(61) |
| Choice of HSP | 49(43) | 65(57) | 23(25) | 69(75) | 24(47) | 27(53) | 96(37) | 161(63) |
| Patient suggesting other friends to come for treatment | 97(85) | 17(15) | 75(82) | 17(18) | 42(82) | 9(18) | 214(83) | 43(17) |
| Patient comes again for treatment | 99(87) | 15(13) | 74(80) | 18(20) | 44(86) | 7(14) | 217(84) | 40(16) |

#### Tangibility

Among the seven basic items of examination room, 95% clients find the doctors room visually attractive. Sphygmomanometer (82%) and stethoscope (77%) was mostly available during examination in all UPHCs. Observation finding showed that stethoscope was available (100%) in all places but sphygmomanometer was not available in counselor and pediatrician room. Availability of thermometer and measuring tape was only around 40% in all UPHC while observation showed that, they were available but used occasionally. Finding showed that, availability of gloves was only 20%, being lowest at KMSS (13%). Our observation finds that, gloves was available in 80% of our observation but HSP did not use gloves during examination in all three UPHCs. Among the clients who had already used the services before, said all the UPHCs’ standard of services was improved (more than 62%), but around 25% of clients said it was not improved and more than 12% clients said it was degrading. Services were degrading more in KMSS and Nari Maitree (15% each).

#### Reliability

Majority of the clients said HSP were reliable when they provide services. UPHCs responded timely (96%) and HSP was dependable (98%). However, in 12% cases, they did not write in OPD records, more in KMSS (22%). Our observation showed that, HSP wrote drug prescriptions in OPD records. However, for ANC checkups HSP wrote examination findings. Full history was not written for the future records.

#### Assurance

In all three UPHCs, HSP gave courage and assurance to clients (96%), there was someone to give assistance (93%) and hence clients felt safe to share their problems (97%). Observation showed, for older physician, courage and assurance was the part of their suggestion whereas for middle aged physician, assurance was not common.

#### Empathy

Out of total responses, 98% said HSP provided dedicated attention during consultation. 91% clients have said that they got enough time to share problems, 96% said HSP were compassionate and 93% clients’ health needs were identified. Consultation time ranges from less than one minutes to 30 minutes with 9 minutes as an average time. It was highest in Nari Maitree (around 10 minutes) and lowest in KMSS (8 minutes). In contrast, our observation showed average consultation time was 5 minutes. (Graph 1)

#### Client satisfaction

According to the service users, rate of services was graded into five categories; poor, unsatisfactory, average, satisfactory and good. Since unsatisfactory and satisfactory had less responses, we merged into poor and average respectively. Quality of services was best in Nari Maitree (75%) followed by BAPSA and KMSS (each 66%). 84% users will go again for treatment and 83% will suggested their friends to go seek services in UPHCs. (Graph 2)

We asked regarding satisfaction in open ended question. In all the UPHCs, clients said that the behavior of HSP, services and management was satisfactory. In multiple answer question, at least 67% clients said behavior of HSP was good in all facilities where more were satisfied in Nari Maitree (88%). Services was satisfactory for 34% clients in both BAPSA and KMSS. But comparatively, management was more satisfactory in Nari Maitree (22%). Here service was experienced doctor, explanation of doctor and free medicine. Management was combined with satisfaction regarding cleanliness, environment and short waiting time. (Graph 3)

#### Client dissatisfaction

Around 15% clients of KMSS said quality was poor but in BAPSA it was only 10%. (Graph 2). Out of 257 responses, 61% did not find all services they wanted in UPHCs. In KMSS, 67% did not find services they wanted where, 59% and 57% were in BAPSA and Nari Maitree. 19% did not get medicines from UPHCs, where KMSS had highest (27%). Around 25% said that, there was no doctor and 63% could not choose service provider. Overall, 61% clients said they did not find the services they wanted.

In all UPHCs, clients described their experiences with services was high waiting time. It was mostly said in BAPSA (77%) and less time said at Nari Maitree (58%). In Nari Maitree, 40% had to explain several times to HSP regarding their problems. But 12% of clients in KMSS and 16% of clients in BAPSA had to ask other HSP about their problems. In KMSS 12% of clients said, HSP did not explain clearly to their clients, where BAPSA and Nari Maitree had only 4% and 0% respectively.

We asked about dissatisfaction of UPHC services. More than 59% clients were dissatisfied with high waiting time in all facilities, highest being in BAPSA (65%). Management was more dissatisfactory for 36% of clients of Nari Maitree. Similarly, behavior of HSP was most dissatisfactory for KMSS (13%). Here, we included “buying medicine from outside”, “HSP are not enough”, “less consultation time”, “services” etc. in “management” category (Graph 4).

#### Waiting time

Average waiting time was 36 minutes where range was from 0 to 200 minutes. Highest average waiting time was in KMSS (41 minutes) where BAPSA had least (38 minutes). However, from our observation, average waiting time of three UPHCs was 61 minutes. (Graph 5)

However, more than 45% of clients in all UPHCs said, their all of their needs were met. It was most clients were satisfied in BAPSA (54%). In Nari Maitree, 45% clients said most of their needs were met which was lowest in KMSS (36%). Around 18% clients of KMSS said their only few needs were met where Nari Maitree was only 4%. Very few (1%) said that, their needs were not met.

## 5. Discussion

Discussion is divided into two parts; responsiveness of HSP and quality of services when provided. Client’s sociodemographic status showed that 82% were from catchment area where almost all clients (99%) were female and married (98%). Most of their age lies between 18 to 24 years. Only, few (10%) were employed.

### Responsiveness of HSP

#### Information

In exit interview, under the information domains, all five individual variables (HSP listening carefully, explain about diseases, patient understanding HSP, facilitation about follow up and patient assessing medical record for future use) were indicating HSP as responsive with 90% score. Similar findings were observed in a study done in rural setting were private HSP were more responsive than public HSP [7, 27]. Our observation using same tool showed that, in the variables of information (patient understanding suggestions was 1.8 and advice for follow up was 2 out of 4), some were below 50% thus findings of exit interview and observations differs. It might be because of either client were satisfied by few information (reflection from exit interview findings) or sample size in our observation was low (49) to give significant result.

#### Friendliness

In friendliness domain, according to result of exit interview, 86% of the respondents reported receiving two ways conversation and socio-cultural friendly conversation from the provider. In rapport building elements (asking client’s name and engaging in social talks), they have only crossed 40% and 70%. Comparatively, HSP of BAPSA had low scores in rapport building. Though studies shows that HSP of private health facilities of rural area were responsive in friendliness domains [7, 8, 20, 28, 29] however in our study overall score of friendliness was observed less than 50% in all categories (rapport building, two way conversation with eye contact, socio-cultural friendly conversation). Studies on client satisfaction reveals that, friendliness behavior of doctor and nurses increased client satisfaction in private hospitals of Dhaka city but comparatively low in public health facilities of same city [30].

Although our research did not compare between private and public, however UPHCs of north city corporations were better than south city corporation due to low patient flow which gives enough time to talk to the clients.

#### Trustworthiness

In gaining trust, clients did not feel HSP was good in providing emergency contact number and service orientation. Comparatively, HSP of KMSS got less score for providing emergency contact number but in terms of service orientation Nari maitree was poor. We did not find any research on providing emergency contact numbers to clients by HSP in Bangladesh.

In exit interview clients of all UPHCs reported HSP misbehave, highest being in BAPSA (26%). According to our observation, though the HSP in all three UPHCs were service oriented but showed minimum respect to patient. Similar study in rural setting showed that, HSP gain more trust in public doctors. In public sector HSP were more sensitive towards financial status of clients [7, 20, 28]. However, in our study, clients were not satisfied with these issues in UPHCs. In other researches done in similar settings showed that, service orientation was the strongest factors to influence clients’ satisfaction. But it was not at satisfactory level in Bangladesh as physician were more oriented towards private hospitals to earn money [30]. Although HSP were service oriented, around 30% client had to pay out of pocket money for the medicine.

#### Non-maleficence

In non-maleficence domain, our finding revealed that people got benefit from treatment with some level of side effect from the treatment. Compare to other centers clients of Nari Maitree mostly reported that they suffer from side effects of prescribed drugs. It might be because HSPs did not ask history in detail or they were not using sterile or new equipment during their procedures. For example, 67% clients were not asked allergic history before prescribing drugs in all facilities which was more common in Nari Maitree and KMSS. Our quality of care findings (Table 4) also suggested that although clients got long consultation time, tangible items or sterile items were not used in many of the consultations in all facilities.

From our study, there was no clear reason behind not using sterile equipment during procedures. Lack of time, lack of motivation and ignorance might be the reasons behind this fact which needs further research to explore.

#### Respect for dignity of person

In this domain, it was good in “interaction with respect” subdomain while it was poor responses in “similar type of services in terms of social status”. BAPSA was not consistent in keeping female attendant for female patient. Finding also showed that, HSPs miss to take consent before examination in all facilities. Comparatively, it was poor in Nari Maitree.

Component such as closing salutation was to be ignored in all the facilities. Despite being an urban healthcare facility, clients were not treated equally in terms of gender, ethnicity, religion, caste, social status and economic status. But more than 80% clients of Nari maitree said services were similar in all social status. However, in our study, 61% clients were from households whose monthly average income ranges from 10000 to 20000 BDT. In other studies also, discrimination was very common in terms of social and economic status in public hospitals [31]. Similar research in rural settings of Bangladesh showed that private doctors give more respect to clients than in public hospitals but HSP were not disrespectful in public as well [7]. Possible reason might be rich people go to seek service in private health facilities [31]. With high patient load and low salary in public hospitals, it could make different in their responses, which can be explored further. In India also, in urban settings, clients expects respect from HSP but get less respect in public hospitals than private hospitals [32].

### Quality of services when provided

#### Tangibility

From our findings from exit interview, some of the basic tangible items (gloves, thermometer etc.) were not available all the times during examination of clients. If it was there, it was not used in all health facilities. WHO indicated that availability and use of tangible items had direct impact in quality of health services. Similar to our findings, studies had shown about 50% items had no use in developing countries [4, 33]. It suggested to emphasize on use of long-term tangible items. Other studies also showed that private health facilities had more tangible items than public facilities [30, 34]. If we assumed UPHCs were a public facility, it cannot be ignored that tangible items were ignored in public institutions.

#### Reliability

Our finding from exit interview showed that, few HSP neglected to write on OPD records, which makes them less reliable. Similar study done in Bangladesh suggested that, people relied more on elder and experienced doctors. Satisfaction and reliability influence each other [30]. Other study suggested that, young doctor did not have interpersonal communication skills, exploring psychosocial history, showing empathy, giving courage and assurance, beginning and ending interviews etc [28, 29]. But our study did not explore on the experience of the doctors. Other factors determining reliability were providing service timely and dependability of HSP, which were more than 98% in all facilities.

#### Assurance

All three UPHCs quality of services were good in terms of assurance as HSP give courage and reassurance to clients and clients also felt safe to share their problems. It was contrast to the other studies done in Dhaka with same study tool which showed that HSP of Bangladesh were not good in terms of assurance [30, 34].

#### Empathy

Clients were satisfied with empathy as more than 90% clients got dedicated attention during consultation, enough consultation time and HSP were able to meet the need of clients. It was in contract to other study done in Dhaka, which showed clients did not get full attention and enough time to consult [30, 34]. From our finding, clients reported average waiting time was 39 minutes and consultation time was around 9 minutes. It was contrast from other studies. In other studies, waiting time ranged from 80 minutes to 150 minutes in different researches [17]. Consultation time ranges from 48 seconds to 2-3 minutes in Bangladesh [35]. But other studies had shown that, consultation time goes up to 6.02 minutes in private hospitals [8, 17, 28, 29, 30, 34]. In our study, most of the clients came for antenatal care check-up. Since it took time to take history and examine abdomen, consultation time came long in our study.

Because of long consultation time, other people had to wait for longer time. Hence, there was long waiting time and consultation time in all three UPHCs.

#### Client Satisfaction

UPHCs did not have system to give option to choose HSP. Many of female clients returned back if they had to encounter male HSP. Moreover, around 60% did not find services they wanted in all UPHCs. Findings also suggested that, even though UPHCs were supplying free medicine, clients were not getting it. Around 30% had to buy medicine from their out of pocket money. However, more than 65% of clients suggested that quality of services was good in all UPHCs, best being Nari Maitree (75%). In all categories (behavior of HSP, service and management), KMSS clients were less satisfied than other health facilities. Most satisfactory thing of three UPHCs was behavior of HSP, maximum being in Nari Maitree.

#### Clients Dissatisfaction

Similarly, most dissatisfactory thing was long waiting time in all facilities. Similar to our findings, other research also suggested that behavior and consultation time had significant relation with client satisfaction [7, 30]. Research done in India suggested that, clients prefer the facilities where they face less waiting time [37]. Other dissatisfactory things from our findings were cleanliness and management of UPHCs. Reason for poor in cleanliness might be scarcity of water in Dhaka city, which can be corrected as pilot study of Dhaka suggested [38].

## 6. Conclusion

Our study concludes that, all UPHC’s HSP gave good informative except knowing whether patient understand or not the suggestions given by HSP during consultation. HSP were very friendly but poor in building rapport, which needs some improvement. Sometimes misbehave by HSP, business-oriented attitude, not providing emergency contact number had reduce trust on HSP. Improper use of tangible items and incomplete history taking had increase side effect during treatment. Though patient expect basic respect from HSP, they were not getting it but rather facing discrimination during provision of services.

Quality of services when provided to clients was very satisfactory in terms of reliability, assurance and empathy domains in all health facilities. Writing on OPD records was poor in KMSS. Use of gloves, thermometer, stethoscope, measuring tape and sterile instrument was very low in three facilities. Some services, medicine, diagnostic tests were not available in UPHCs. People had to pay out of pocket money to buy medicine and services. Clients were mostly satisfied with behavior of doctor. Services and management of UPHC satisfied them in some instance. Long waiting time was the most dissatisfactory experience for clients. Comparatively clients were highly satisfied with the services of Nari Maitree.

## 7. Recommendation

There are some places to improve the services. Some of the areas are discussed here.

- Number of HSP should be increased as per population coverage to ensure sufficient time for quality consultation, with options to choose male or female provider.
- Training on behavior change communication for HSP including paramedics, cleaners, office assistance should be initiated to increase clients’ satisfaction.
- Clarity on clients understanding of instruction given during consultation should be ensured by HSP.
- An emergency contact number should be provided from the center to clients. That would be more useful for pregnant women [39].
- HSP must ensure proper procedure (e.g. use of sterile tangible items, taking detail history on OPD records) to reduce side effects from procedures. Clients consent should be obtained before procedure.
- Waiting time should be minimized by managing patients (e.g. making order/time table of patients).
- UPHC should find ways to reduce out of pocket expenditure for medicine and services ensuring delivery of all the required essential drugs timely.
- Overall environment of UPHC should be improved be clean and healthy by improving management UPHC, motivation of cleaners and strict governance in all UPHCs.
- Scope of expanding service package to include If it is feasible with the budget other services like nutrition, endemic diseases, adolescent health of primary health care should be emphasized to improve utilization of UPHCs.
- Though services for geriatrics and non-communicable diseases were enlisted in the service package, it was not provided actively [13].

## 8. Strength and Limitation

We used two data collection method (exit interview and observation) and triangulated it to find the similarities to capture as much as variability in responses. With the time constrains and feasibility of the study, we have some limitation in our study. We had planned to take two UPHCs from each DNCC and DSCC but since one UPHC took more time to give permission, we had to manage with one UPHC from South City Corporation. Since our sample size was calculated to be 384, we could collect only 257. In UPHC, their services were focused to both gender but as community believe that the service were only for female, we were bound to select 98% of female in our exit interviews. During analysis, since we planned to analyses by multivariate methods, due to limited resources to do exploratory factorial analysis, we only presented descriptive analysis in our study. Since observation and exit interview clients were not same and were conducted in different time, criterion validity could not be ensured.

## Data Availability

All relevant data are within the manuscript and its Supporting Information files. For further contact

## 9. Acknowledgement

I would like to offer my special thanks to my supervisors Prof. Dr. Syed Masud Ahmed and Mr. Kuhel Faizul Islam, JPGSPH, BRAC University, who guided me through out my summative learninig project. I also acknowledge my mentors Dr. Nahitun Naher and Ms. Roksana Haque, JPGSPH, BRAC University for their continuous support and guidance from the beginning of this project. I am grateful to the supports provided by Dr. Purabi Ahmed, BAPSA, Md Tauhidul Islam, KMSS and Rehana Akhter Mita, Nari Maitree who permitted us to do our study in their respective UPHCs. I am also thankful to Mr. Kousik Ahmed and the MPH team for logistic and technical support during SLP. At last but not least, I am thankful to my group mates and research assistant who supported me during this project.

## Annex 1: Operational Definition

Responsiveness of HSP: It is defined as the activities which meet social, ethical and human rights of clients when services are being delivered at UPHC by HSP to meet genuine expectation of clients (Murray and Frenk, 1999; WHO, 2000).

**Quality of health care when provided:** It is defined as the services which meets minimum standards of care, safe services, affordable to the society, reliable, and ability to improve health outcomes which finally meets client expectation and satisfy them, when service is being delivered by HSP in UPHC (USAID, 2004).

**Access:** It refers to means of approaching UPHC to utilize the available primary health care services.

**Utilization:** It is defined as the use of available primary health care services of UPHC by the beneficiary.

**Beneficiary:** It refers to a person who receives health services or advice from health service provider in UPHC. Is also known as patient or client.

**Variables of responsiveness variables;**

**Information:** It is delivering understandable messages by HSP to the beneficiary to explain them about the procedure, medicines, consequences of treatment, and clarify all the queries they have. Friendliness: A quality of HSP to be friendly with clients during service delivery.

**Trustworthiness:** It refers to become trustful towards clients by gaining trust in the treatment that HSP provides.

**Beneficence:** It refers to proper treatment following standards to provide medical, social and economic benefit to clients without harm to the clients.

**Respect for dignity of person:** It refers to the respect shown by HSP to beneficiary by protecting from potential abusive practices, physical, and mental harms to ensure rights and freedoms of clients.

**Variables of quality of services when provided;**

**Technical competence:** It refers to services provided by the technical and skilled HSP.

**Reliable:** It is ability of HSP to perform promised services accurately.

**Assurance:** Ability of HSP to give confidence towards the services of UPHC during service provision.

**Empathy:** Ability of HSP to understand clients’ problems and feelings during service delivery.

**Client satisfaction:** It refers to achievement of the quality of services as per the expectation and needs of clients while delivering services in UPHC.

**Graph 1.**
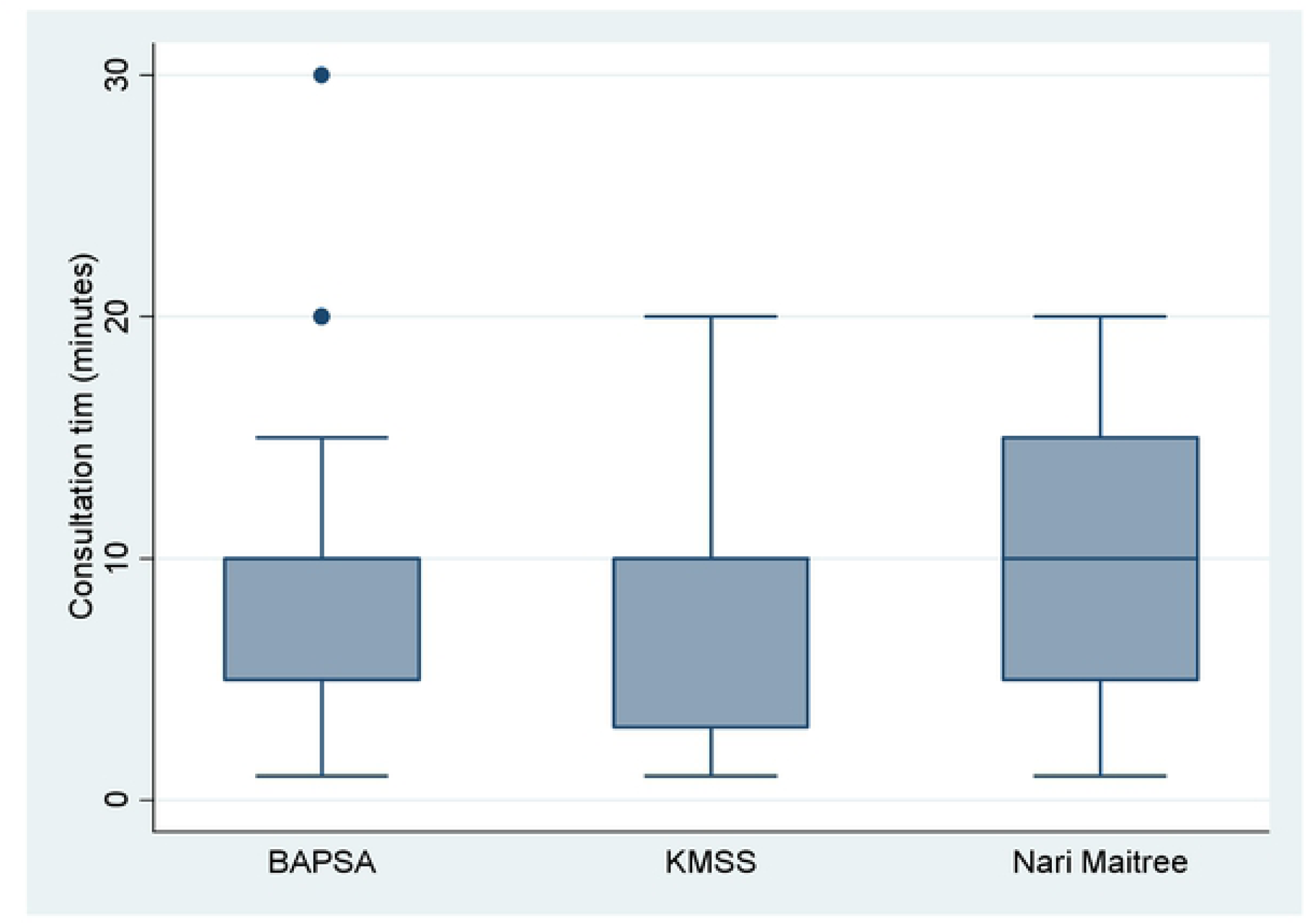
Box plot showing consultation time of three UPHCs

## Annex 2: Questionnaire tools

**Accessibility to and utilization of Urban Primary Health Care**

**This tool is divided into two parts: Interviews and Observation**

**Date of Interview: _____________ Time:_______________ Place: ____________________**

### I. Interviews

**A. Socio-demographic profile**

| S.N. | Questions | Response |
| --- | --- | --- |
| 1. | Name of the respondent _____ |  |
| 2. | Sex of the respondent | 1. Male<br>2. Female<br>3. Other |
| 3. | Age of respondent in years<br>What is your child's age in months/years<br>(below 18) |  |
| 4. | Marital status of respondent | 1. Single<br>2. Married<br>3. Separated<br>4. Divorced<br>5. Widowed |
| 5. | Religious affiliation of respondent | 1. Islam<br>2. Hindu<br>3. Christian<br>4. Buddhist<br>5. Other |
| 6. | How many children do you have? |  |
| 7. | How many family members do you have including you? _____ |  |
| 8. | What is your present address?<br>Since when you are living here? _____ | 1. City corporation (specify)<br>.....<br>2. District town (specify)<br>.....<br>3. Other town<br>(specify).....<br>4. Village (specify)<br>..... |
| 9. | What is your permanent address? | 1. City corporation (specify) .....<br>2. District town (specify) .....<br>3. Other town (specify) .....<br>4. Village (specify)<br>..... |
| 10. | If you are not a permanent resident of this area, what was the main reason for moving to the current place? | 1. Looking for work<br>2. More earning<br>3. Own education<br>4. Children's education<br>5. Familial<br>6. Marriage<br>7. Buy new land/house<br>8. Look after properties<br>9. River erosion<br>10. Eviction<br>11. Security<br>12. Other (specify) ..... |
| 11. | Do you come from ____ wards (the catchment area) of UPHC? | 1. Yes<br>2. No |
| 12. | What is your educational level? | 1. Illiterate<br>2. Primary<br>3. Secondary<br>4. Higher Secondary<br>5. Graduation<br>6. Post-graduation<br>7. No formal education<br>8. Technical training |
| 13. | Are you employed...? | 1. Yes 2. No , **if no go to 16** |
| 14. | Employment status | 1. Full time<br>2. Part time |
| 15. | What is your occupation? | 1. Service (Specify.....<br>2. Small business<br>3. Large business<br>4. Farmer<br>5. Teacher<br>6. Others..... |
| 16. | What is your average monthly household income? |  |
| 17. | Does your household possess these goods? | 1. Refrigerator<br>2. TV<br>3. Mobile phone<br>4. Radio<br>5. Motor bike<br>6. Sewing machine<br>7. Bicycle |
|  |  | 8. IPS<br>9. Motor car |
| 18. | What is your source of drinking water? | 1. Wasa supply tap in home<br>2. Tube well<br>3. Wasa street tap<br>4. Pond<br>5. Deep tube well<br>6. Others..... |
| 19. | What is the process of water purification you practice? | 1. Boiling<br>2. Filter<br>3. Water purification tablet<br>4. Nothing<br>5. Others..... |
| 20. | How is your household infrastructure made of? | 1. Brick build<br>2. Kachha<br>3. Tin shed |
| 21. | What type of toilet do you use? | 1. Sanitary latrine<br>2. Drain<br>3. Pond<br>4. Hanging toilet |
| 22. | From where do you take treatment if you are usually ill? | 1. Traditional healer<br>2. Pharmacy<br>3. UPHC<br>4. Government hospital<br>5. Private hospital |
| 23. | What is the distance of the UPHC from your home? | ____kilometers/walking distance |
| 24. | What time usually needed to reach to the UPHC? | 1. 10-15 min<br>2. 30min<br>3. 1hour<br>5. More than 1 hour |
| 25. | What transport do you need to reach to UPHC? | 1. By walk<br>2. Rickshaw<br>3. CNG<br>4. Bus<br>5. Car |
| 26. | What is the usual transport cost needed to reach UPHC? | 1. 10 to 20tk<br>2. 50tk<br>3. 100tk<br>4. More than 100 tk |
| 27. | Is this your first visit/ follow up visit?<br>Skip to 32, if first visit | 1. First visit(if 1, then go to 32)<br>2. Follow up |
| 28. | How many times did you visit the health facility in last three months | 1. 1-2 times<br>2. 3-4 times<br>3. More than 4 times<br>4. Cannot remember |
| 29. | Why did you visit/what was your/your child's problem? What was the purpose of visit? |  |
| 30. | Was your problem being solved in this UPHC center? | 1. Yes<br>2. No<br>3. Referred to other place |
| 31. | What is the purpose of today's visit (your /your child's problem, the day of interview)?<br>(Which department do you want to go?) |  |
| 32. | Has the physician given any diagnostic tests for your disease? | 1. Yes<br>2. No |
| 33. | Has the physician given any advice for your disease? | 1. Yes<br>2. No |
| 34. | Do you know what are the services available in the health center..... | 1. Yes<br>2. No (if no skip to 37) |
| 35. | Can you name the services available in this UPHC. |  |
| 36. | Do you know the prices of services....? | 1. Yes (If no. go to 40)<br>2. No |
| 37. | Can you mention the service prices. |  |
| 38. | Are these services reasonable in price/costly? | 1. Yes<br>2. No<br>3. Costly |
| 39. | Do you get medicines from this UPHC? | 1. Yes<br>2. No |
| 40. | Are these medicines free? | 1. Yes<br>2. No |
| 41. | Do you buy medicines from outside (Private pharmacy)? | 1. Yes<br>2. No |
| 42. | How did you manage the entire cost of your health problem | 1. Out of pocket<br>2. loan<br>3. free |
|  |  | 4. Others |

**B. Exit Interview with service users**

**I. Responsiveness of Health Service Provider (HSP)**

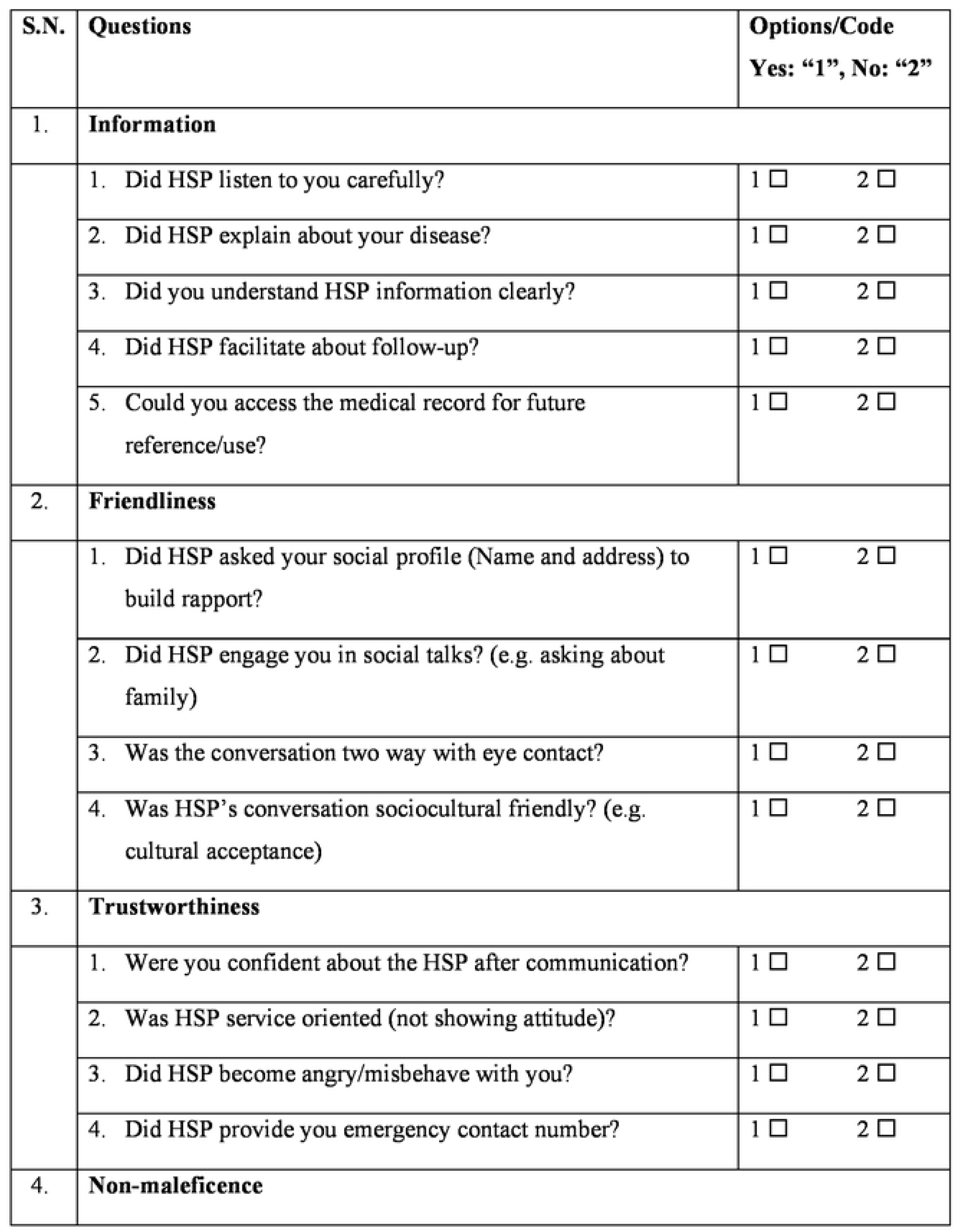
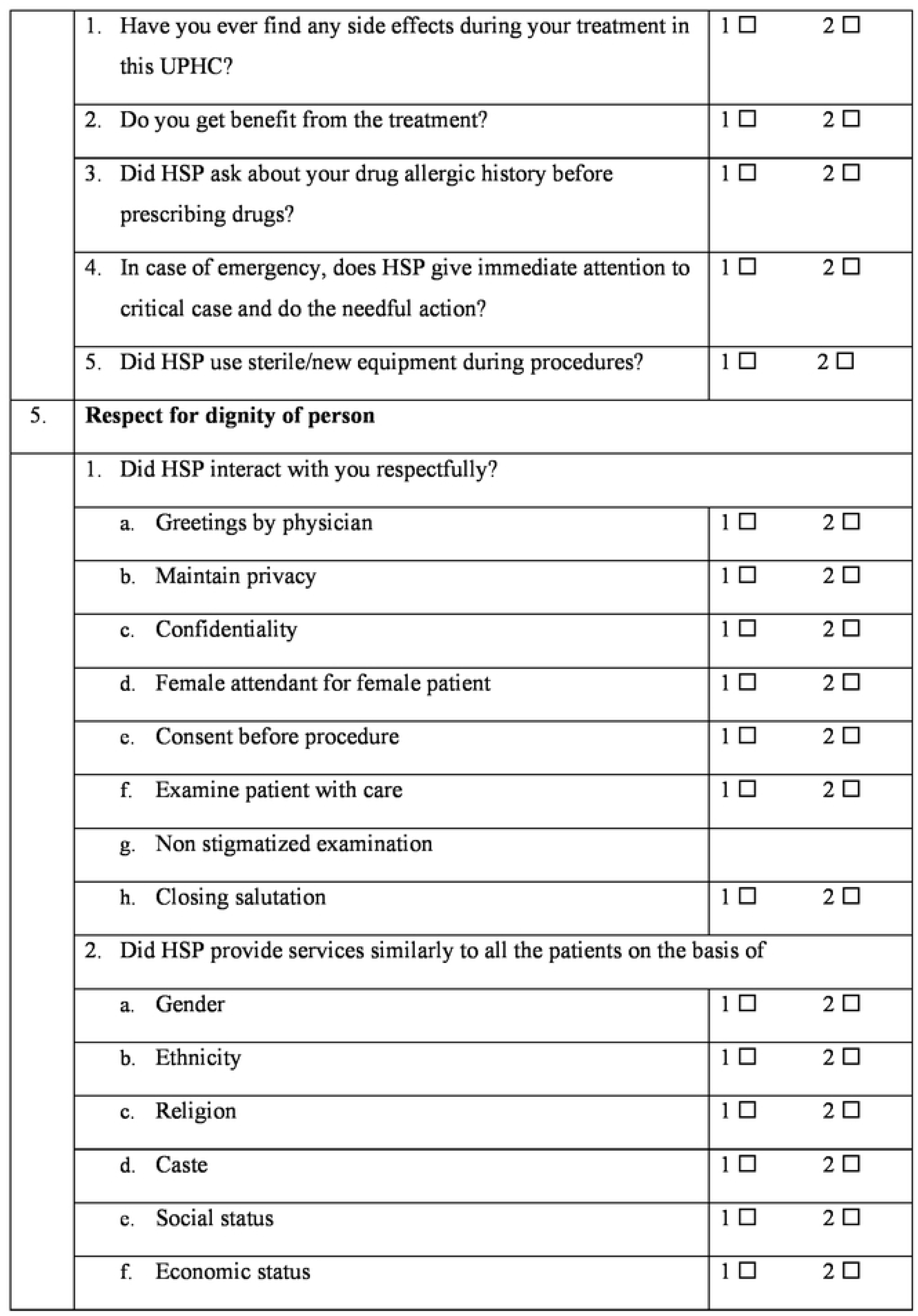

**II. Quality of services when provided**

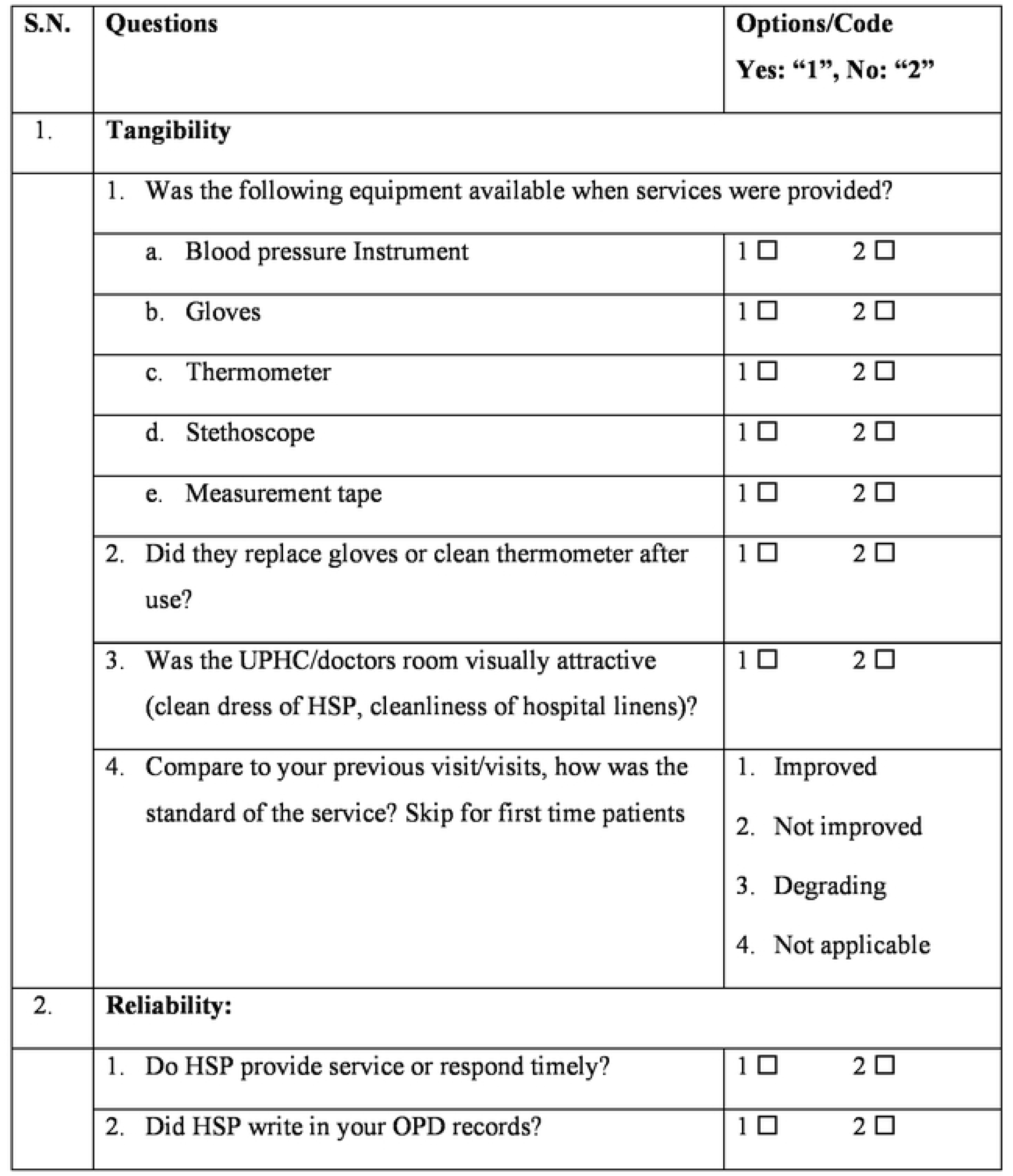
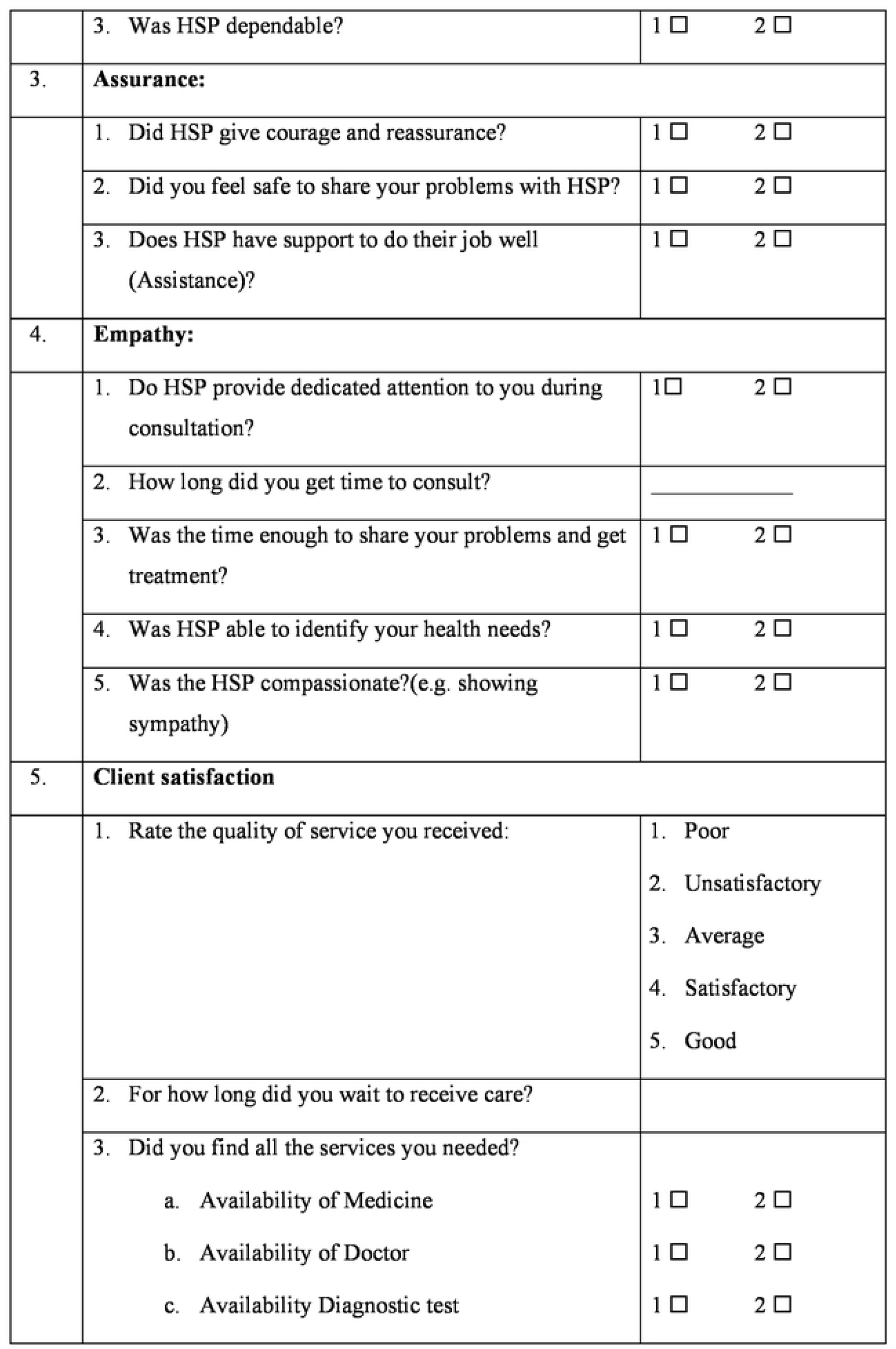
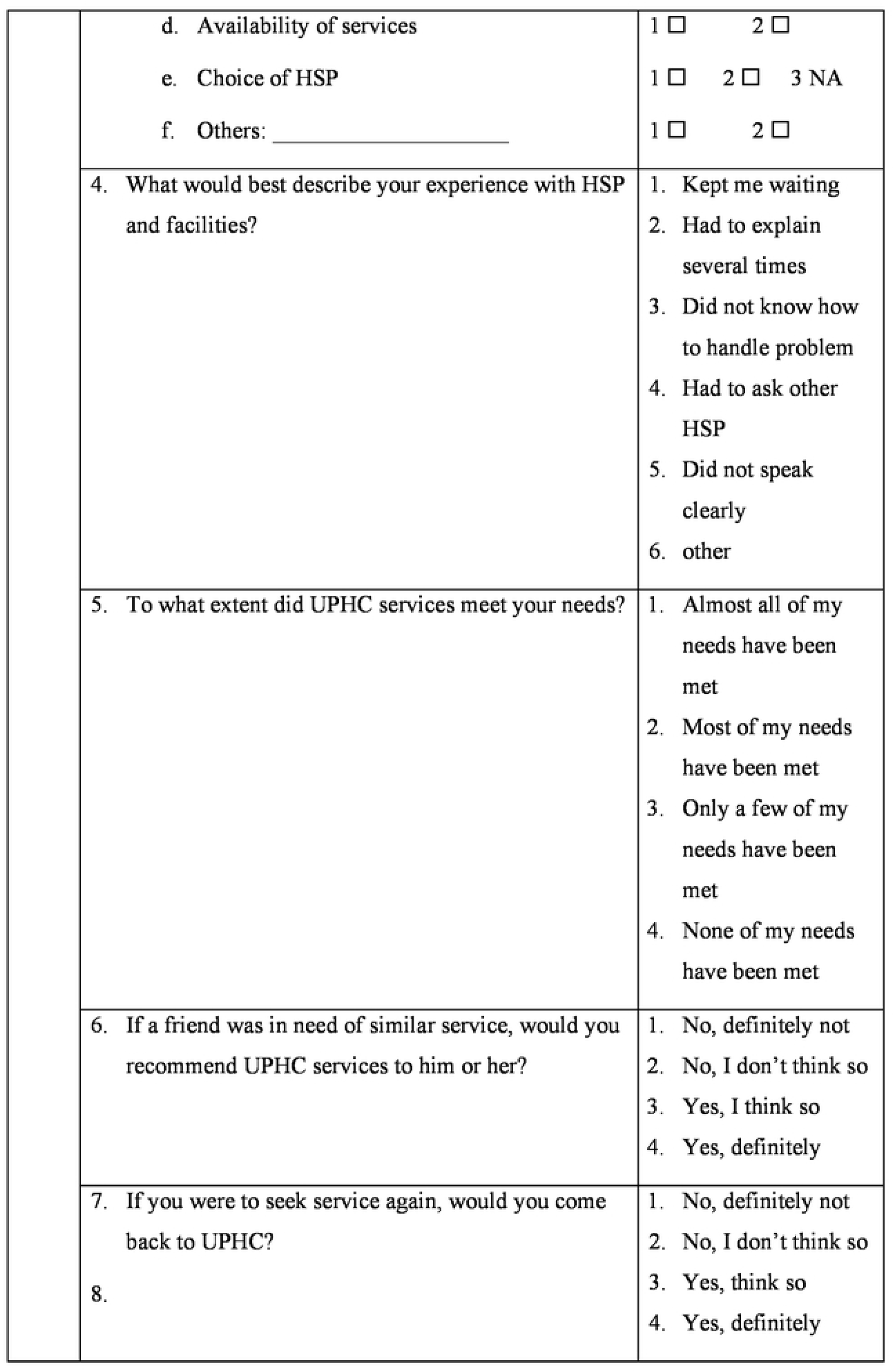
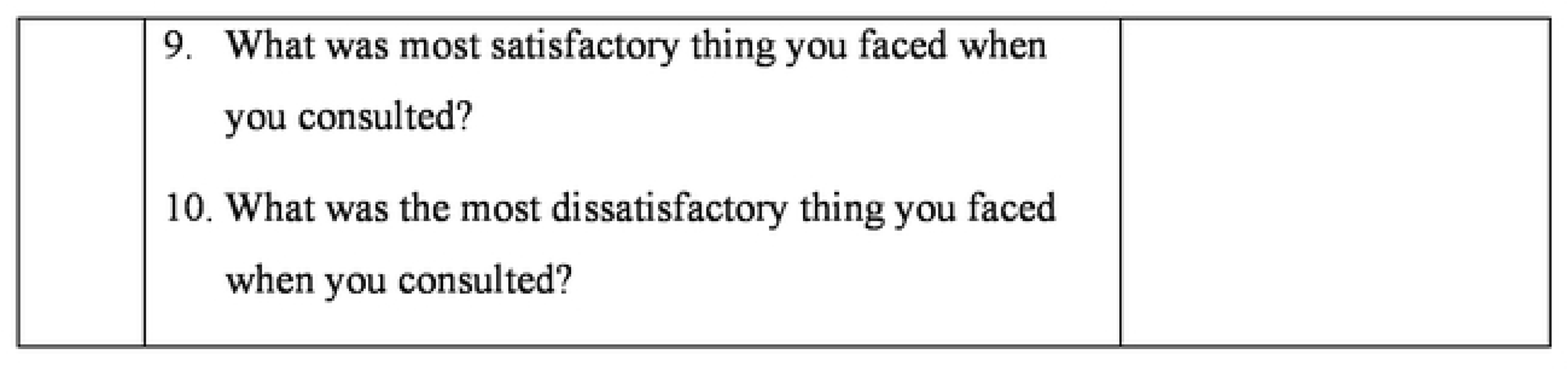

| S.N. | Questions | Options/Code<br>Yes: "1", No: "2" |
| --- | --- | --- |
| 1. | **Tangibility** |  |
|  | 1. Was the following equipment available when services were provided? |  |
|  | a. Blood pressure Instrument | 1 ☐ 2 ☐ |
|  | b. Gloves | 1 ☐ 2 ☐ |
|  | c. Thermometer | 1 ☐ 2 ☐ |
|  | d. Stethoscope | 1 ☐ 2 ☐ |
|  | e. Measurement tape | 1 ☐ 2 ☐ |
|  | 2. Did they replace gloves or clean thermometer after use? | 1 ☐ 2 ☐ |
|  | 3. Was the UPHC/doctors room visually attractive (clean dress of HSP, cleanliness of hospital linens)? | 1 ☐ 2 ☐ |
|  | 4. Compare to your previous visit/visits, how was the standard of the service? Skip for first time patients | 1. Improved<br>2. Not improved<br>3. Degrading<br>4. Not applicable |
| 2. | **Reliability:** |  |
|  | 1. Do HSP provide service or respond timely? | 1 ☐ 2 ☐ |
|  | 2. Did HSP write in your OPD records? | 1 ☐ 2 ☐ |
|  | 3. Was HSP dependable? | 1 ☐ 2 ☐ |
| 3. | **Assurance:** |  |
|  | 1. Did HSP give courage and reassurance? | 1 ☐ 2 ☐ |
|  | 2. Did you feel safe to share your problems with HSP? | 1 ☐ 2 ☐ |
|  | 3. Does HSP have support to do their job well (Assistance)? | 1 ☐ 2 ☐ |
| 4. | **Empathy:** |  |
|  | 1. Do HSP provide dedicated attention to you during consultation? | 1 ☐ 2 ☐ |
|  | 2. How long did you get time to consult? | _____ |
|  | 3. Was the time enough to share your problems and get treatment? | 1 ☐ 2 ☐ |
|  | 4. Was HSP able to identify your health needs? | 1 ☐ 2 ☐ |
|  | 5. Was the HSP compassionate?(e.g. showing sympathy) | 1 ☐ 2 ☐ |
| 5. | **Client satisfaction** |  |
|  | 1. Rate the quality of service you received: | 1. Poor<br>2. Unsatisfactory<br>3. Average<br>4. Satisfactory<br>5. Good |
|  | 2. For how long did you wait to receive care? |  |
|  | 3. Did you find all the services you needed? |  |
|  | a. Availability of Medicine<br>b. Availability of Doctor<br>c. Availability Diagnostic test | 1 ☐ 2 ☐<br>1 ☐ 2 ☐<br>1 ☐ 2 ☐ |
|  | d. Availability of services<br>e. Choice of HSP<br>f. Others: _____ | 1 ☐ 2 ☐<br>1 ☐ 2 ☐ 3 NA<br>1 ☐ 2 ☐ |
|  | 4. What would best describe your experience with HSP and facilities? | 1. Kept me waiting<br>2. Had to explain several times<br>3. Did not know how to handle problem<br>4. Had to ask other HSP<br>5. Did not speak clearly<br>6. other |
|  | 5. To what extent did UPHC services meet your needs? | 1. Almost all of my needs have been met<br>2. Most of my needs have been met<br>3. Only a few of my needs have been met<br>4. None of my needs have been met |
|  | 6. If a friend was in need of similar service, would you recommend UPHC services to him or her? | 1. No, definitely not<br>2. No, I don't think so<br>3. Yes, I think so<br>4. Yes, definitely |
|  | 7. If you were to seek service again, would you come back to UPHC?<br>8. | 1. No, definitely not<br>2. No, I don't think so<br>3. Yes, think so<br>4. Yes, definitely |
|  | 9. What was most satisfactory thing you faced when you consulted?<br>10. What was the most dissatisfactory thing you faced when you consulted? |  |

**C. Observation Tool**

**ROP-Scale in the form of Structured Observation Tool**

**Understanding and Measuring Responsiveness of Health Service Provider (HSP) in UPHC in Dhaka City**

1. General Identification Questions

Instruction to the observer: Fill out these information’s just before starting the interview with the HSP and structured observation.

2. Observation ID _________________________

3. Date of observation ______________________

4. Name of UPHC __________________________

5. Address of UPHC _________________________

6. Type of provider: Exclusively UPHC/ both UPHC and private

7. Starting time of observation ________________________

8. Ending time of observation ___________________________

Questions for the HSP

**Instruction to the observer:**

9. ID of the HSP _________________________

10. Gender _____________________________

11. Degrees [E.g., MBBS, FCPS (Medicine)] _________________________

12. Number of months in practice ______________________________

13. Number of months working in this UPHC ________________________

14. Average patient examination per day ___________________________

**Observation Items**

**Instruction to the observer:** Be present while consulting with 4,h patients. First 3rd observations will not be recorded. Assess the role of the HSP in this regard by 1 to 4 where 1-completely unsatisfactory and 4-completely satisfactory. Following observation will be graded according to this scale.

**Beginning part**

15. **Greet and welcome to the patient by HSP** and make the patient feel comfort.

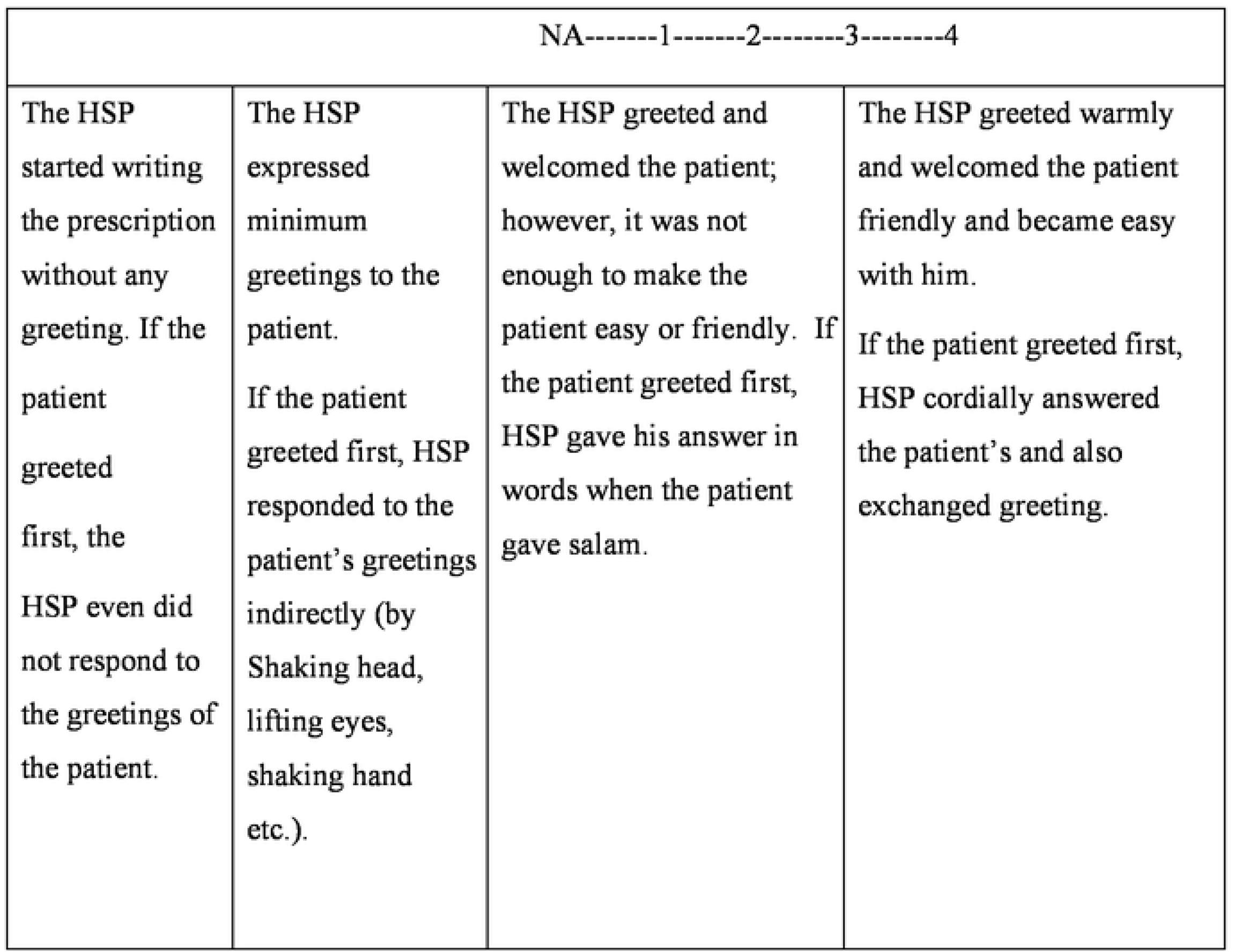

| NA-----1-----2-----3-----4 |  |  |  |
| --- | --- | --- | --- |
| The HSP started writing the prescription without any greeting. If the patient greeted first, the HSP even did not respond to the greetings of the patient. | The HSP expressed minimum greetings to the patient.<br>If the patient greeted first, HSP responded to the patient's greetings indirectly (by Shaking head, lifting eyes, shaking hand etc.). | The HSP greeted and welcomed the patient; however, it was not enough to make the patient easy or friendly. If the patient greeted first, HSP gave his answer in words when the patient gave salam. | The HSP greeted warmly and welcomed the patient friendly and became easy with him.<br>If the patient greeted first, HSP cordially answered the patient's and also exchanged greeting. |

16. Patients expect that the **HSP will ask patient’s name** at least and will treat the patient as a person.

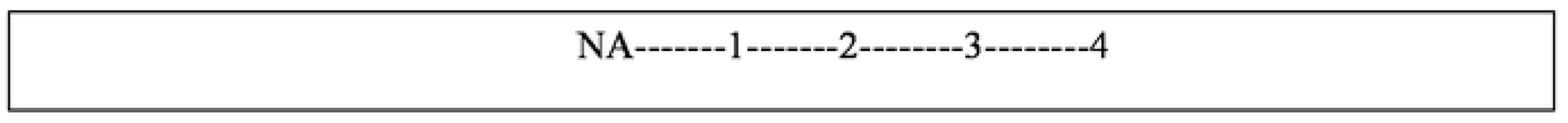
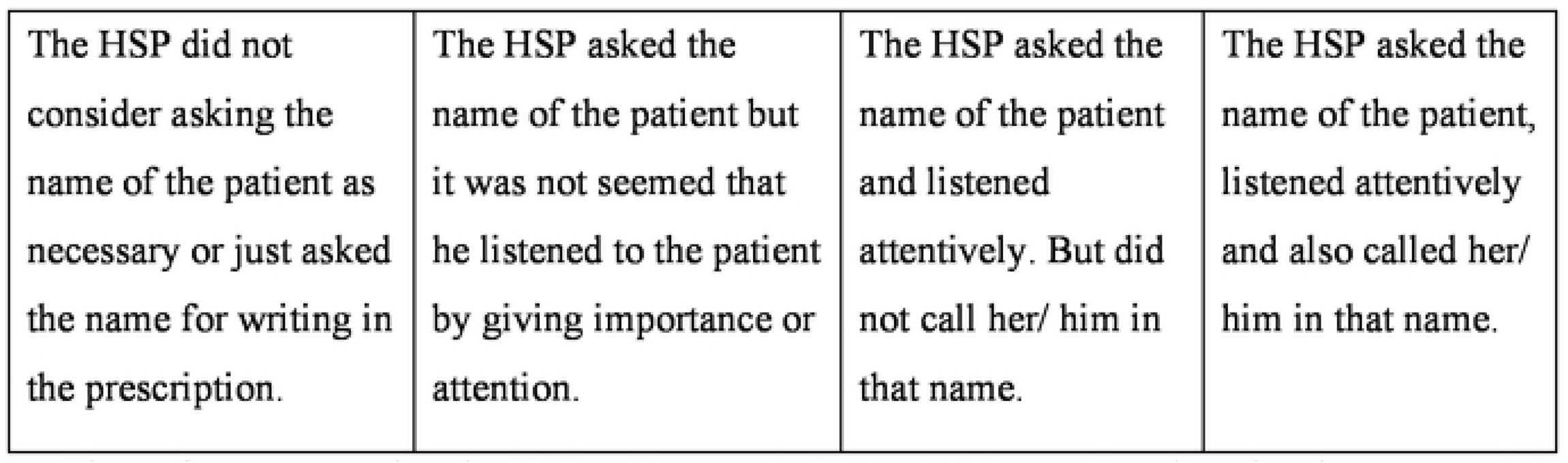

|  |  |  |  |
| --- | --- | --- | --- |
| The HSP did not consider asking the name of the patient as necessary or just asked the name for writing in the prescription. | The HSP asked the name of the patient but it was not seemed that he listened to the patient by giving importance or attention. | The HSP asked the name of the patient and listened attentively. But did not call her/ him in that name. | The HSP asked the name of the patient, listened attentively and also called her/ him in that name. |

17. The patient expects that the **HSP will not only listen to his problem** but also do some **social talks** and **listen to the patient** if he does any social talk.

| NA-----1-----2-----3-----4 |  |  |  |
| --- | --- | --- | --- |
| The HSP did not do any social talk with patient. Even he stopped the patient when the patient started to do some social talks. | The HSP did minimum social talk or responded when the patient started social talk. | The HSP did some social talk or responded when the patient started social talk. | The HSP got involved completely into social talk or if the patient started, he participated satisfactorily into social talks. |

18. The patient expects that HSP will also **ask about his family**.

| NA-----1-----2-----3-----4 |  |  |  |
| --- | --- | --- | --- |
| The HSP did not ask anything about the family of the patient or there was | The HSP did not want to know anything about the family of the patient but there were some talking regarding this in his advice. | The HSP wanted to know about the family of the patient but not in details. That was not a part of the treatment rather a part of social talk. | The HSP wanted to know about the family of the patient in details cordially. That was not a part of the treatment rather a part of social talk. |
| nothing in this regard during his consultation. | That means it was a part of his treatment but not a social talk. | **For example:** how many members are there in his family, how many children, are they all well etc. | **For example:** how many members are there in his family, how many children, are they all well etc. |

19. Patients expect that the **HSP will be friendly**.

|  |  |  |  |
| --- | --- | --- | --- |
| NA-----1-----2-----3-----4 |  |  |  |
| The HSP was not friendly. | The HSP was friendly at a minimum level. | The HSP was somewhat friendly. | The HSP was fully friendly. |
| The example of friendliness may be: remembering the name and face of the patient and calling him by name (here 'calling by name' means calling by name of the patient in a friendly manner); asking or making comment about an event of the patient's family; praising the patient (about clothing or anything else); asking for an opinion of the patient about anything (weather, politics etc.) |  |  |  |

20. Patients do not **expect misbehave** rather **expect good behavior** from the HSP.

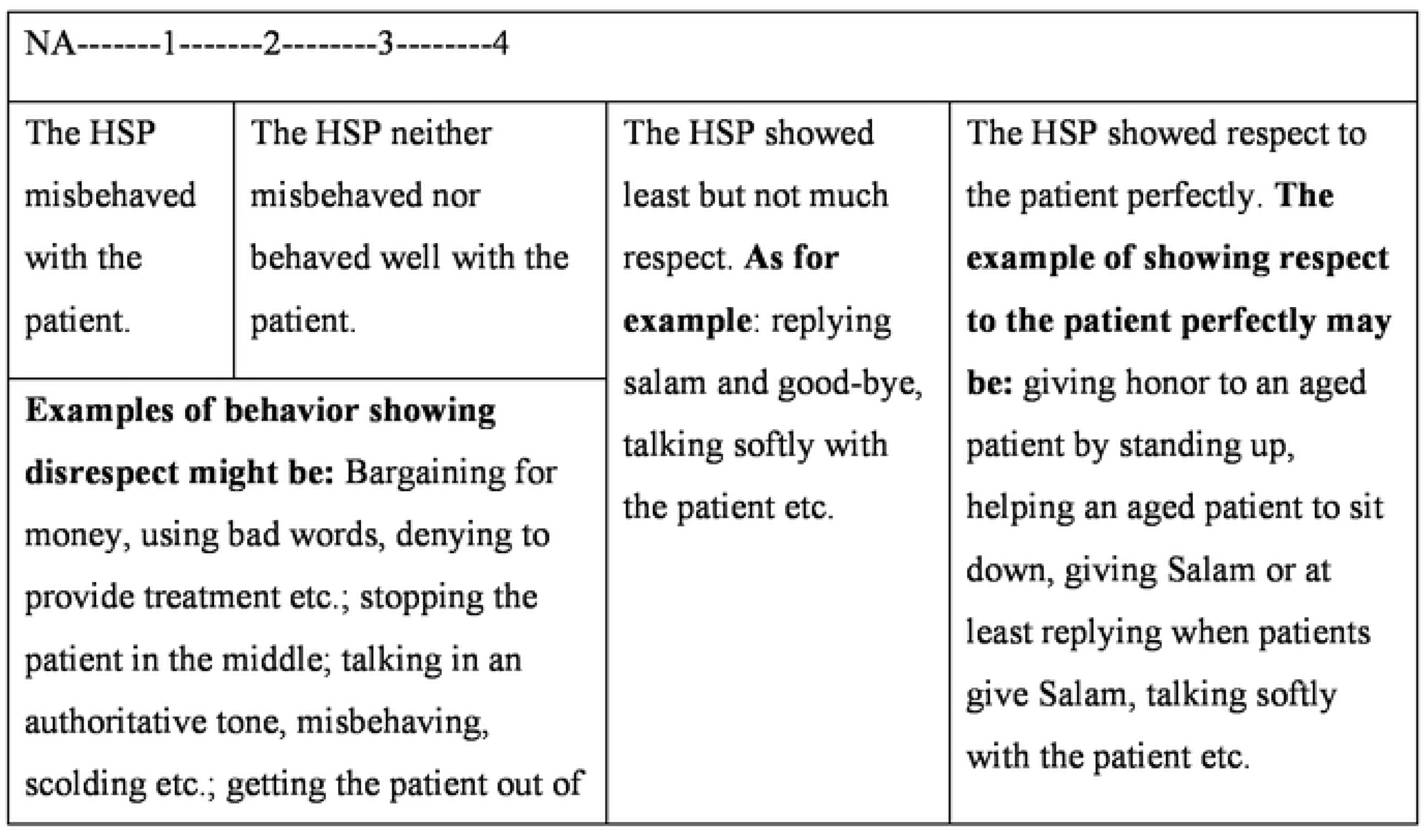
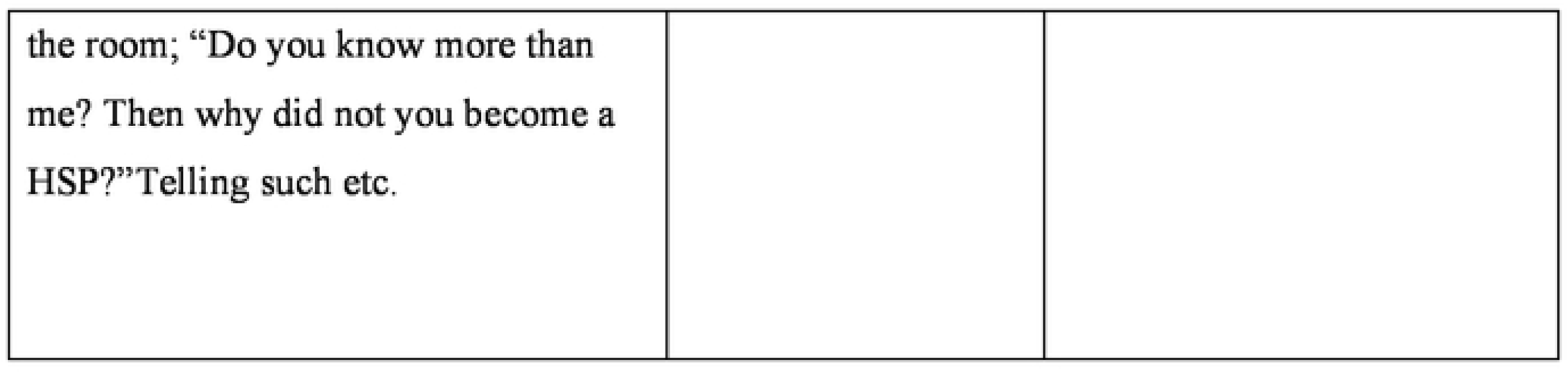

|  |  |  |  |
| --- | --- | --- | --- |
| NA-----1-----2-----3-----4 |  |  |  |
| The HSP misbehaved with the patient. | The HSP neither misbehaved nor behaved well with the patient. | The HSP showed least but not much respect. **As for example:** replying salam and good-bye, talking softly with the patient etc. | The HSP showed respect to the patient perfectly. **The example of showing respect to the patient perfectly may be:** giving honor to an aged patient by standing up, helping an aged patient to sit down, giving Salam or at least replying when patients give Salam, talking softly with the patient etc. |
| **Examples of behavior showing disrespect might be:** Bargaining for money, using bad words, denying to provide treatment etc.; stopping the patient in the middle; talking in an authoritative tone, misbehaving, scolding etc.; getting the patient out of |  |  |  |

|  |
| --- |
| the room; “Do you know more than me? Then why did not you become a HSP?” Telling such etc. |

**History Taking**

21. Patients expect that the HSPs will **start writing the prescription after listening to the symptoms** in detail and completely.

| NA-----1-----2-----3-----<br>---4 |  |  |  |
| --- | --- | --- | --- |
| The HSP did not listen to the patient attentively. The HSP stopped the patient while describing his complains. | The HSP listened to some of the patient's complaints, but at some stage HSP stopped the patient either by verbal or non-verbal cues. HSP had gone to the next step (examining, prescribing, etc.) before the patient finished describing his complaints. | The HSP listened to patient's complaints, did not stop the patient in the middle. But HSP had gone to the next step (examining, prescribing, etc.) before the patient finished describing his complaints. | The HSP heard the patient up to the end, did not interrupt (except necessary questions). He started next step only when the patient completed the description of the disease in details. |

22. Patients expect that the **HSP will listen to them attentively** with patience and that would be expressed by his behavior.

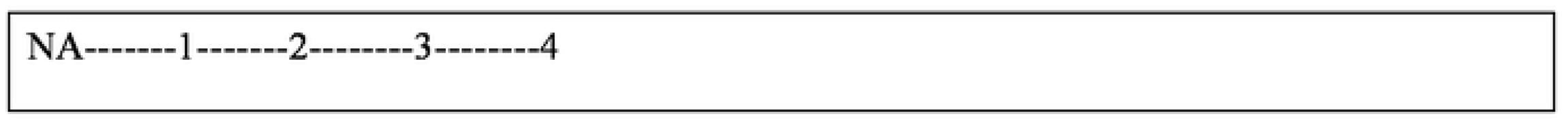
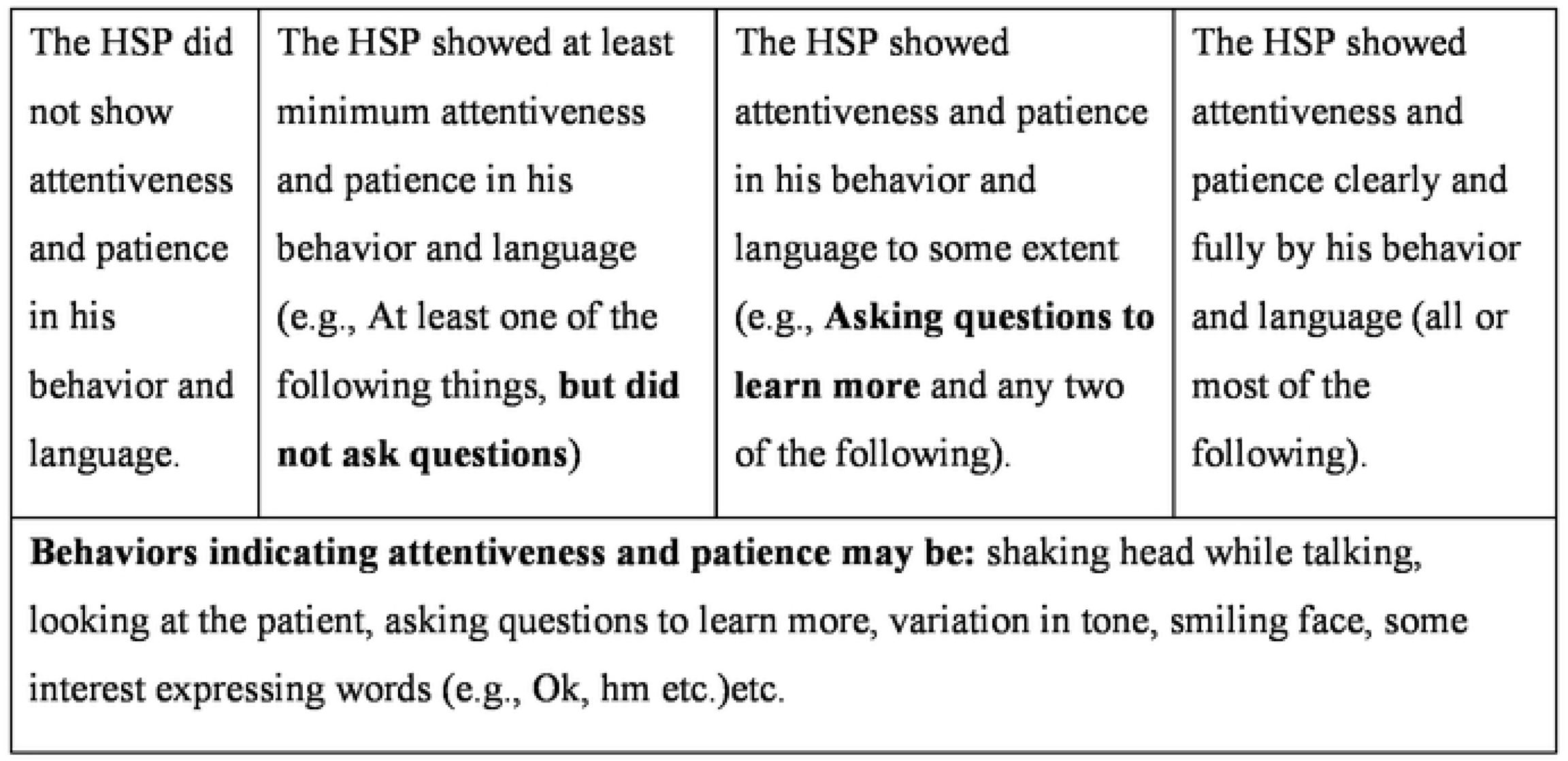

|  |
| --- |
| NA-----1-----2-----3-----4 |

|  |  |  |  |
| --- | --- | --- | --- |
| The HSP did not show attentiveness and patience in his behavior and language. | The HSP showed at least minimum attentiveness and patience in his behavior and language (e.g., At least one of the following things, **but did not ask questions**) | The HSP showed attentiveness and patience in his behavior and language to some extent (e.g., **Asking questions to learn more** and any two of the following). | The HSP showed attentiveness and patience clearly and fully by his behavior and language (all or most of the following). |
| **Behaviors indicating attentiveness and patience may be:** shaking head while talking, looking at the patient, asking questions to learn more, variation in tone, smiling face, some interest expressing words (e.g., Ok, hm etc.)etc. |  |  |  |

**Examination**

23. Patients expect that the HSP would do the **necessary physical examination with care**.

|  |  |  |  |
| --- | --- | --- | --- |
| NA-----1-----2-----3-----4 |  |  |  |
| The HSP did not examine the patient any more. | The HSP examined the patient at least once (only measured body temperature or pulse rate etc.). | The HSP measured the body temperature, blood pressure, pulse rate etc. (which were necessary) with some care. | The HSP measured the body temperature, blood pressure, pulse rate etc. and did all with care. |
| The example of examining the patient physically may be: telling the patient politely to fold up his sleeves, telling the patient that what he is going to do etc. |  |  |  |

**Prescription Writing**

24. Patients expect that the HSP would not only treat the disease but also **suggest some preventive measures**.

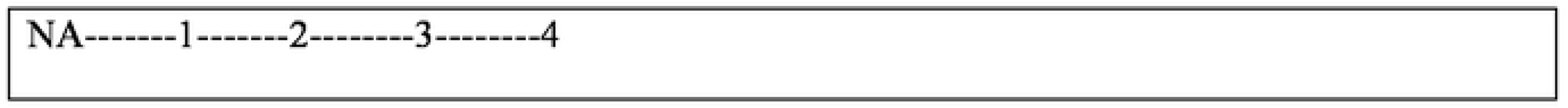
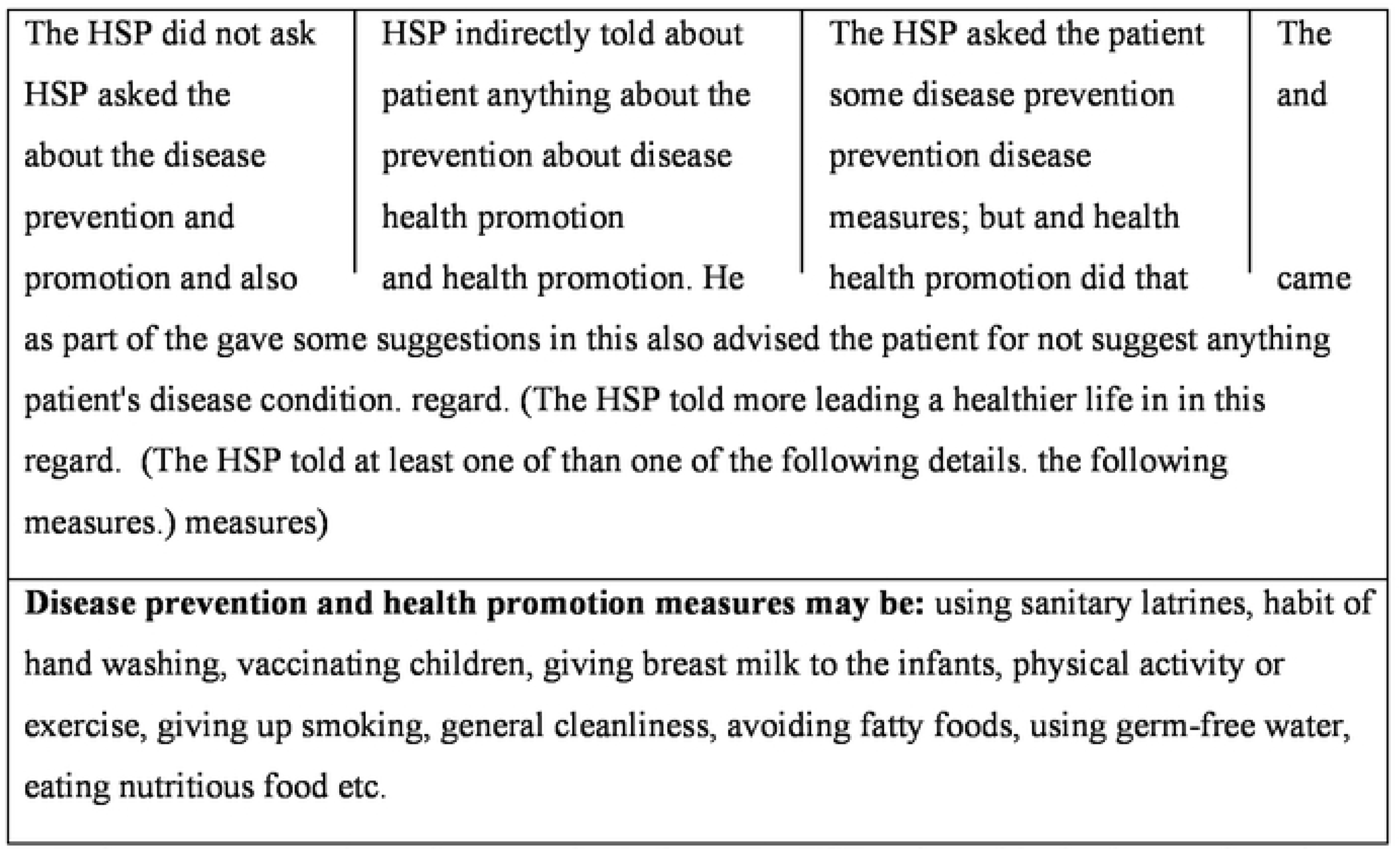

25. Patients expect **service-oriented attitude from the HSP** and consider business-oriented attitude as unwanted.

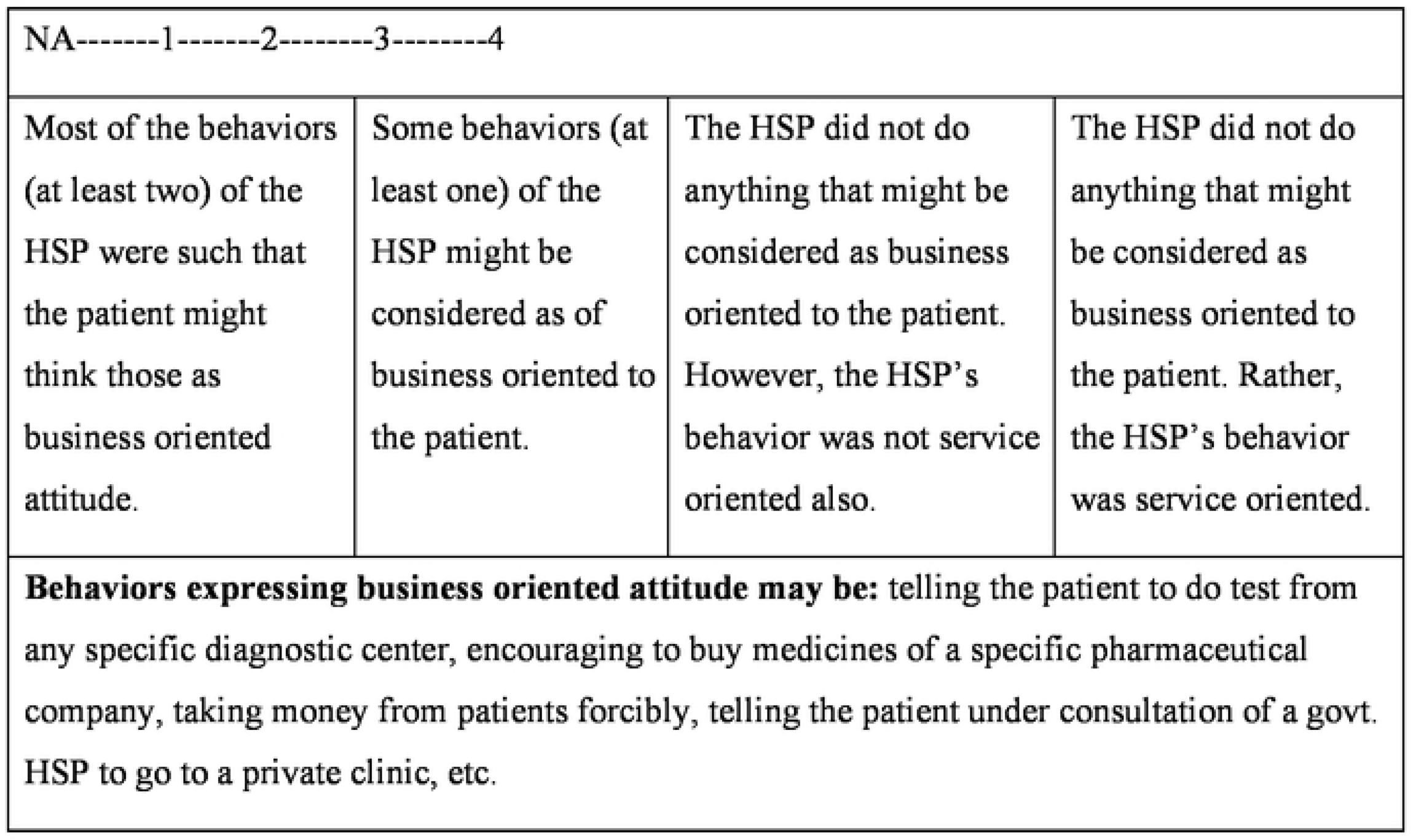
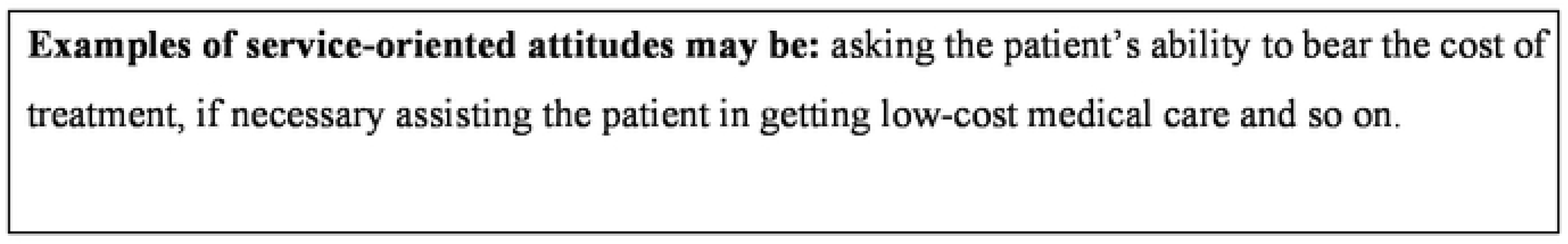

26. Patients expect that the **HSPs would facilitate post treatment follow-up** and give them a follow-up plan.

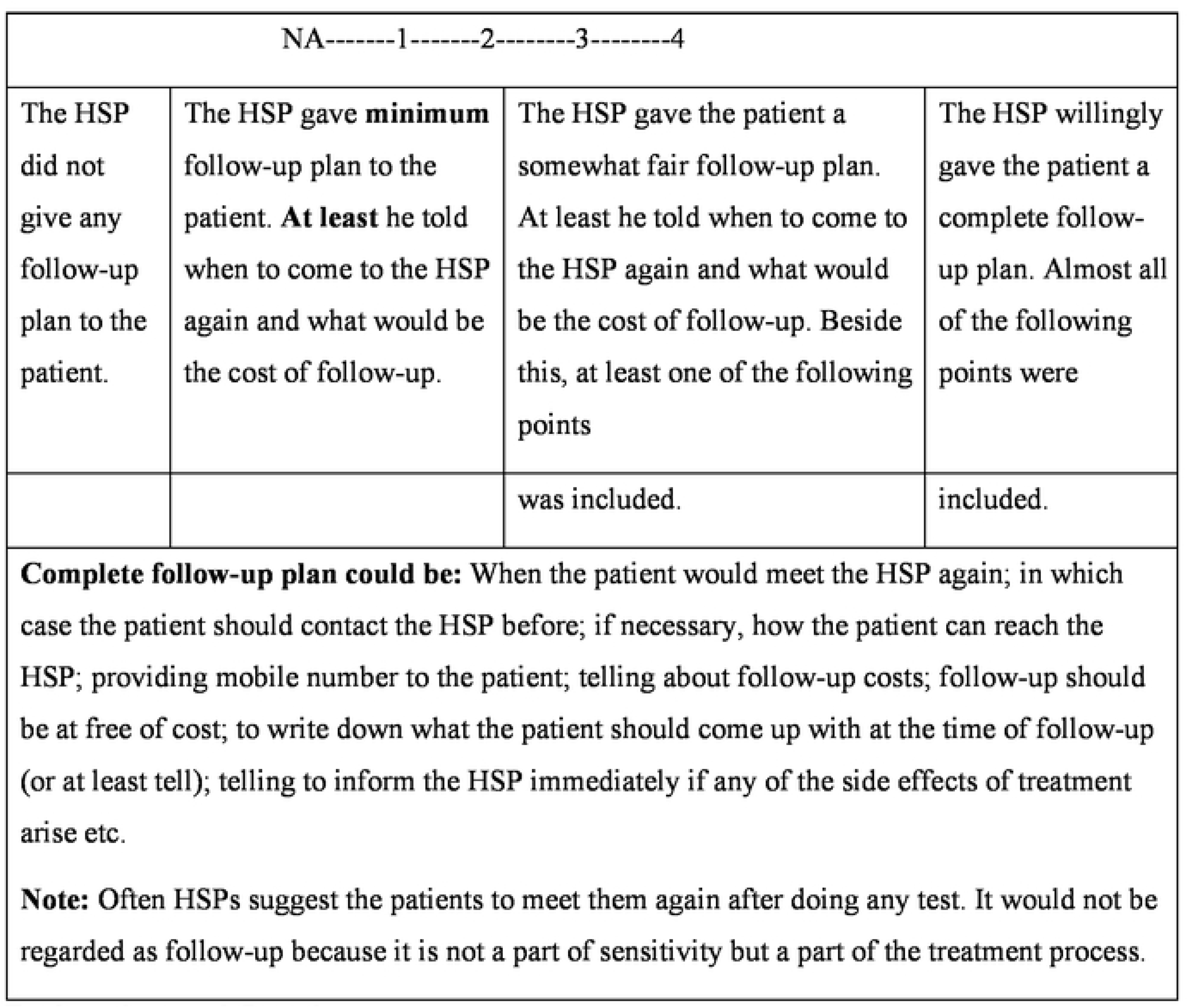

| NA-----1-----2-----3-----4 |  |  |  |
| --- | --- | --- | --- |
| The HSP did not give any follow-up plan to the patient. | The HSP gave **minimum** follow-up plan to the patient. **At least** he told when to come to the HSP again and what would be the cost of follow-up. | The HSP gave the patient a somewhat fair follow-up plan. At least he told when to come to the HSP again and what would be the cost of follow-up. Beside this, at least one of the following points | The HSP willingly gave the patient a complete follow-up plan. Almost all of the following points were |
|  |  | was included. | included. |
| **Complete follow-up plan could be:** When the patient would meet the HSP again; in which case the patient should contact the HSP before; if necessary, how the patient can reach the HSP; providing mobile number to the patient; telling about follow-up costs; follow-up should be at free of cost; to write down what the patient should come up with at the time of follow-up (or at least tell); telling to inform the HSP immediately if any of the side effects of treatment arise etc.<br>**Note:** Often HSPs suggest the patients to meet them again after doing any test. It would not be regarded as follow-up because it is not a part of sensitivity but a part of the treatment process. |  |  |  |

**Explanations and Questions**

27. Patients expect the HSP would **explain everything to them**; cause of the disease, diagnosis, prognosis and severity, treatment, the side effects of the drugs, the result of diagnostic tests (if any), preventive measures of disease (Diet) etc.

| NA-----1-----2-----3-----4 |  |  |  |
| --- | --- | --- | --- |
| The HSP did not explain anything to the patient. He only wrote the prescription. | The HSP explained at least one thing to the patient. Such as:<br>diagnosis (at least the name of the disease), treatment (at least explaining prescription) etc. | The HSP explained many issues to the patient. Such as: diagnosis (at least the name of the disease), treatment (at least explaining prescription), the result of diagnostic tests (if any), preventive measures of disease (Diet) etc. | The HSP explained everything to the patient. Such as: cause of the disease, diagnosis (at least the name of the disease), treatment (at least explaining prescription), the side effects of the drugs (if any), the result of diagnostic tests (if any), preventive measures of disease (Diet) etc. |

28. It is extremely important that the **patient understands all suggestions** or explanations given by the HSP. The HSP should be sure that the patient understands him.

| NA-----1-----2-----3-----4 |  |  |  |
| --- | --- | --- | --- |
| The HSP did not ask the patient about his understanding after explaining him the cause of the disease, diagnosis (name of the disease), | The HSP asked the patient about his understanding after explaining him at least one (especially name of the disease and/or treatment) of the following issues, such as, cause of the disease, diagnosis (name of the disease), prognosis, treatment etc. | The HSP asked the patient about his understanding after explaining him at least two or more of the following issues, such as, cause of the disease, diagnosis (name of the disease), | The HSP asked the patient about his understanding after explaining him each of the following issues, such as, cause of the disease, diagnosis (name of the disease), |
| prognosis,<br>treatment etc. |  | prognosis, treatment<br>etc. | prognosis, treatment<br>etc. |

29. Some of the **behaviour of the HSP discourages patients to ask questions**.

| NA-----1-----2-----3-----<br>4 |  |  |  |
| --- | --- | --- | --- |
| The patients were greatly discouraged to ask questions by the words or behavior of the HSP.<br>**Examples of discouraging behavior may be:**<br>repeatedly looking at the clock, giving reminder to the patient to be short, writing prescription while answering the question, ultraseriousness, answering very shortly (in one word), answering in nagging tone, remaining many patients together at the queue, "you understand more," or "you are the HSP" - telling such neglectful words etc. | The patients might be slightly discouraged to ask questions by the words or behavior of the HSP.<br>**Examples of such behavior could be:**<br>repeatedly looking at the clock, writing prescription while answering the question, answering very shortly (in one word), waiting many patients at the queue, etc. | The words or behavior of the HSP did not discourage patients to ask question. However, the HSP also did not behave in such way to provide courage or to encourage the patients for asking questions. **Example of encouraging conduct may be:**<br>smile answered, carefully listening to the questions, etc. | The words or behavior of the HSP did not discourage patients to ask question; rather the HSP behaved in such way that provided courage or encouraged the patients for asking questions. **Example of encouraging conduct may be:**<br>smile answered, carefully listening to the questions, etc. |

30. One of the most important impediments for patients to understand HSP’s **advice is the medical terminology** (jargon), professional language etc. So, the HSP should avoid such language or explain it if used. [If the HSP does not say any word to the patient, fill in the ‘Not Applicable’ field.]

| NA-----1-----2-----3-----4 |  |  |  |
| --- | --- | --- | --- |
| The HSP used many medical terminologies, and did not explain it to the patient. | The HSP used many medical terminologies, and explained some of those to the patient. | The HSP used one or two medical terminologies, and also explained. | The HSP did not use any medical terminology. |
| **Example of medical terms:** Pus cell (puj), absorb (digestion), nebulize (providing gas) |  |  |  |

**Leaving**

31. **Saying good-bye** is also as important part of consultation as greetings. In this regard, HSP should tell the **summary of consultation**, verify whether the patient has understood everything properly, tell the patient how to communicate with the HSP if any problem arises later and say goodbye with smile.

| NA-----1-----2-----3-----4 |  |  |  |
| --- | --- | --- | --- |
| The HSP did not say good-bye; even when patient said goodbye he did not reply. | The HSP hardly said good-bye to the patient. When patient said goodbye to the HSP, he replied by using gesture (by nutation, eye contact, waving hand etc.). | The HSP said good-bye to the patient but it was not complete or enough. When patient said goodbye, the HSP replied him verbally. **Examples may be:** saying walaikumassalam; ok, you may go; be well etc. | The HSP said goodbye to the patient properly. When patients said good-bye, the HSP responded warmly. **As a good practice, following issues might be included:** summarizing the whole consultation; verifying whether the patient has understood everything properly or not; telling the patient how to communicate with the HSP if any problem arises later and saying goodbye with smile (assalamualaikum, khodahafez, be well, etc.) |

**Throughout Consultation**

32. Patients do not want to see the **HSP involved in illegal or unethical activity**, especially if it is related with their treatment. The HSP should remain away from such type of activity.

| NA-----1-----2-----3-----4 |  |  |  |
| --- | --- | --- | --- |
| The HSP was involved with various illegal activities. | The HSP was involved with at least one illegal activity. | The HSP was not directly involved with any illegal activity. However, such kind of activities happened in front of him and he remained silence that means he had tacit support. Examples: ignoring presence of brokers (inside the room), behavior with medical representatives seen to be over treated etc. | The HSP was not involved with any illegal activity and he was not seen to support any such activities either directly or indirectly. |
| **Examples of such illegal activities may be:** Taking money from patients against free services; bringing patients with the help of brokers; presence of brokers around the chamber (HSPs did not take any step to stop this); collusion with diagnostic centers. Accepting gift from medical representative (and prescribing medicine of that company); taking advantage from brokers (utilizing in hospital, utilizing for personal work) and so on. |  |  |  |

33. Patients expect that the HSP would have **sense of humor** and would provide treatment by becoming easy with patients using his sense of humor.

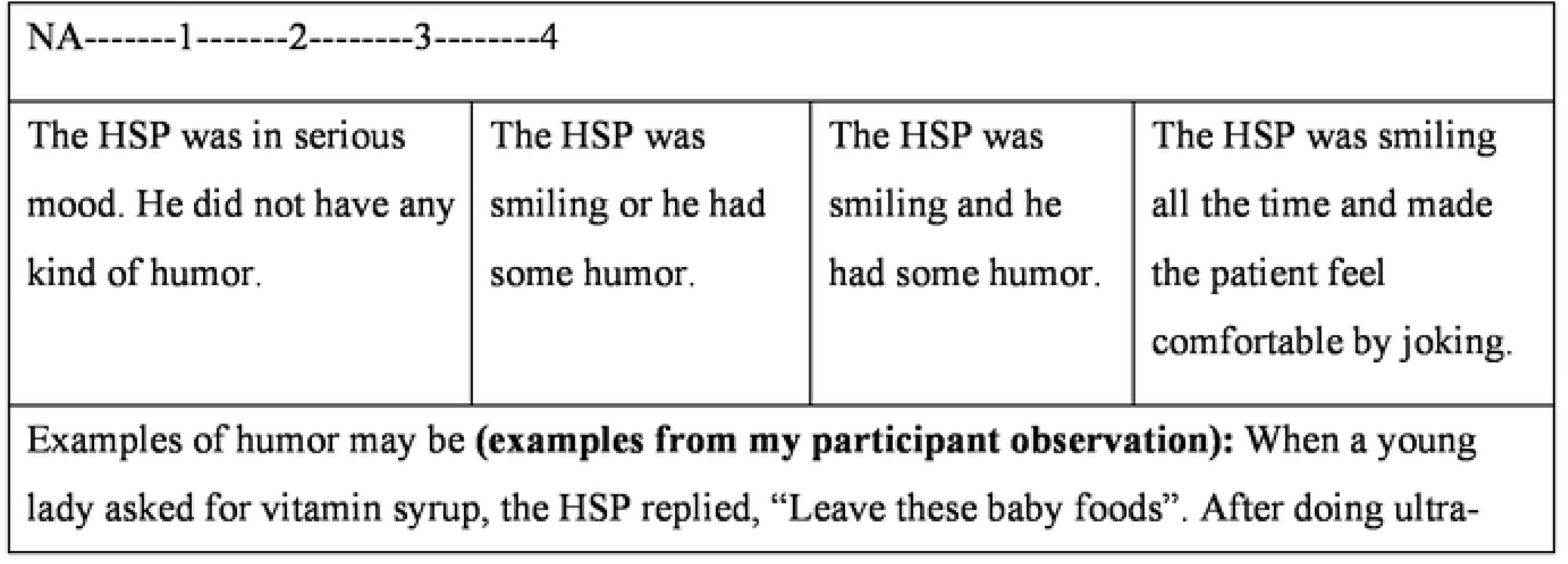
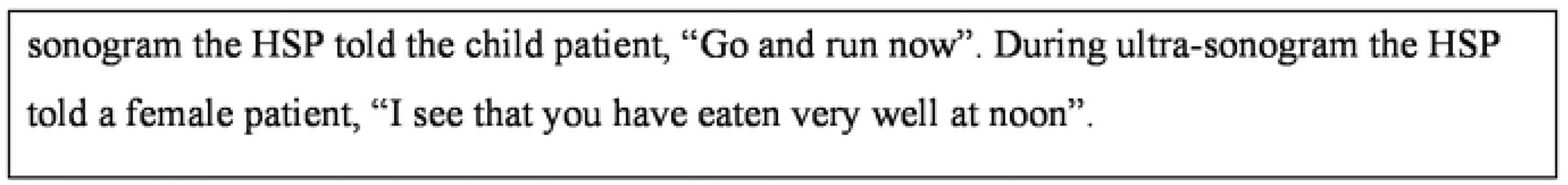

34. Record the consultation time (in seconds) using a stop watch. _____________

35. Age of the patient _______________

36. Gender of the patient ____________________

**Graph 2.**
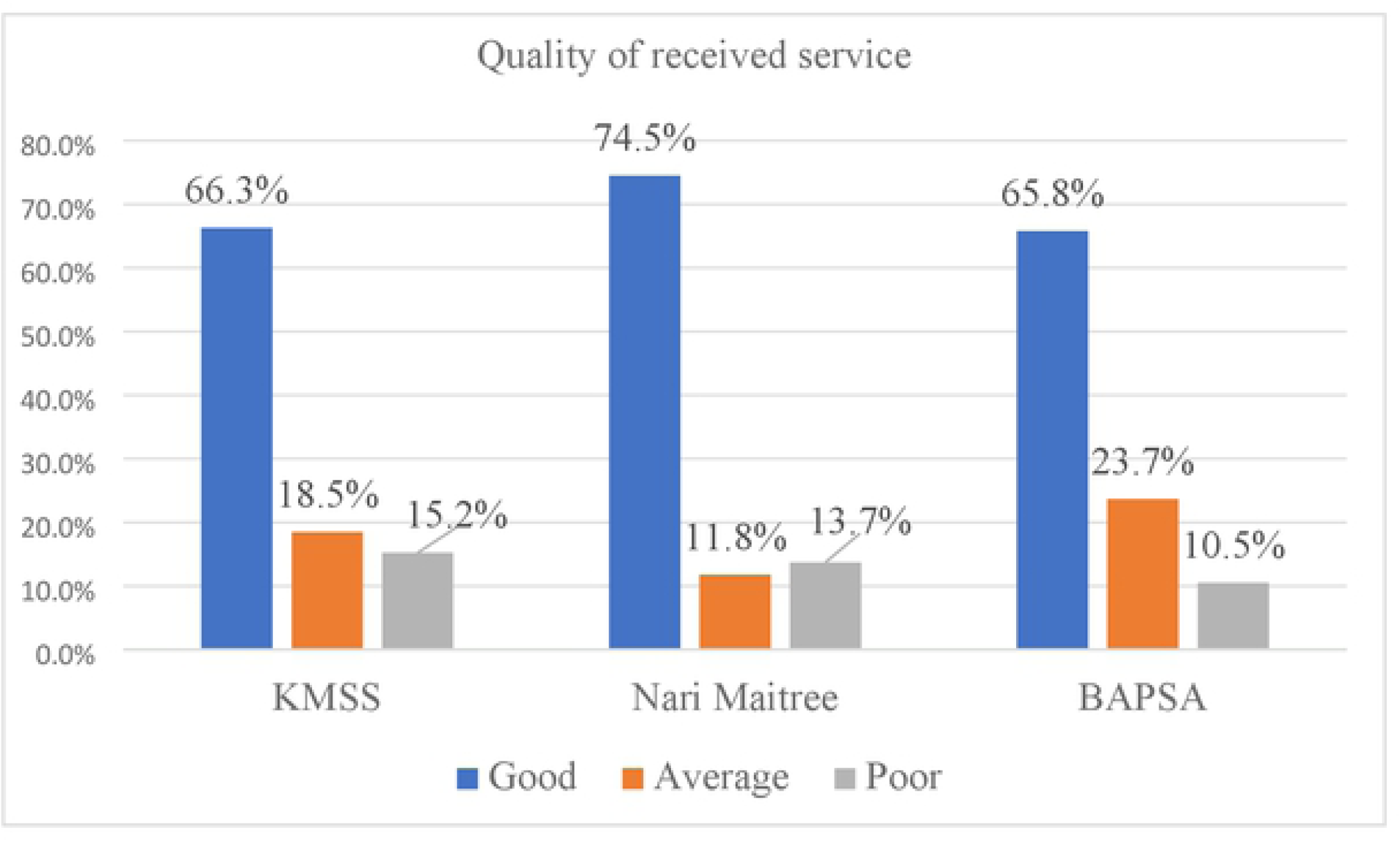
Quality of services (according to users) in three health facilities

## Annex 3: Consent form

### Information and Consent Form for Exit interview

Serial no: Date:

**Date:**
|  |  |
| --- | --- |
| Interview Topic | Accessibility to and Utilization of Urban Primary Health Care Services in Dhaka City |
| Name of the group members | Abhiyan Gautam, Bandana Bhandari, Chungano Hassan, Fatiha Sultana, Saima Mehjabeen |
| Name of the Institution | James P Grant School of Public Health, BRAC University, Bangladesh |

Greetings,

We are students of 13th batch of MPH programme at James P. Grant School of Public Health, BRAC University. As a part of our summative learning project (study activities), we are conducting a study on accessibility and utilization of urban primary health care services. Objective of the study is to explore the current status, enabling factors, challenges towards accessibility and utilization of urban primary healthcare services in Dhaka city. To obtain the study objective we need to interview both the users, caretakers of the users (if age is below-18 years) and providers of UPHC services of this centre. Hence, we are interested to talk with you. We would like to take your permission before proceeding. Your valuable information is really important to us and will lead to better insights and knowledge about the access and utilization of UPHC in Dhaka city. The interview will take 25-30 minutes. There is no direct benefit for you in this study and you will not receive any monitory compensation or gifts. Your responses will remain confidential and anonymous and will be used for purpose of the study only. The information collected from the study will be kept covert by the research group. In order to best understand, we need to ask some questions regarding access and utilization of UPHC services. Keep in mind that your participation will be completely voluntarily. You can withdraw from the interview∕discussion at any moment if you want to, even after signing the consent or beginning the interview. Moreover, you are not obliged to answer any question that makes you feel uncomfortable. There are no restrictions and risks of answering our questions.

Would you be willing to participate in the study?

If yes, please give your signature below

I have read the foregoing information, or it has been read to me. I have had the opportunity to ask questions about it and any questions I have been asked have been answered to my satisfaction. I consent voluntarily to be a participant in this study

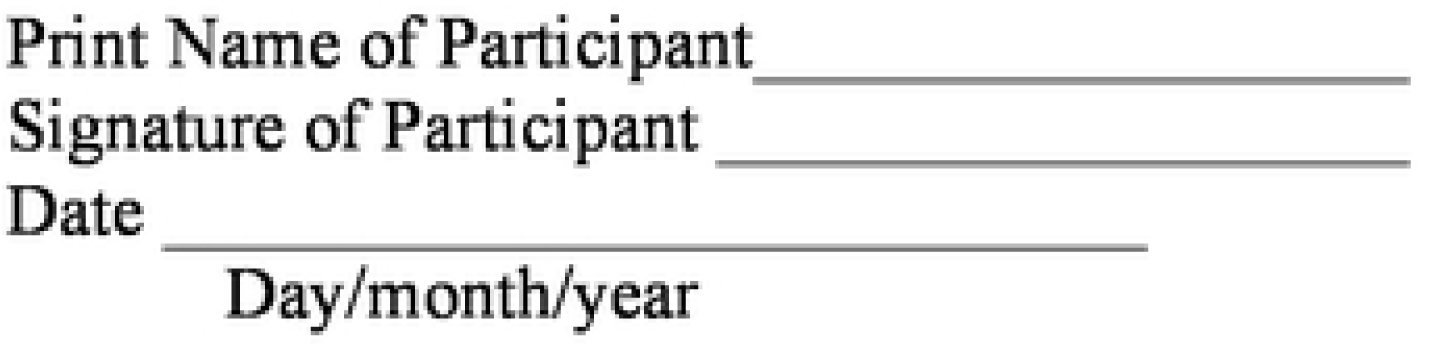

*If illiterate^*^*

I have witnessed the accurate reading of the consent form to the potential participant, and the individual has had the opportunity to ask questions. I confirm that the individual has given consent freely.

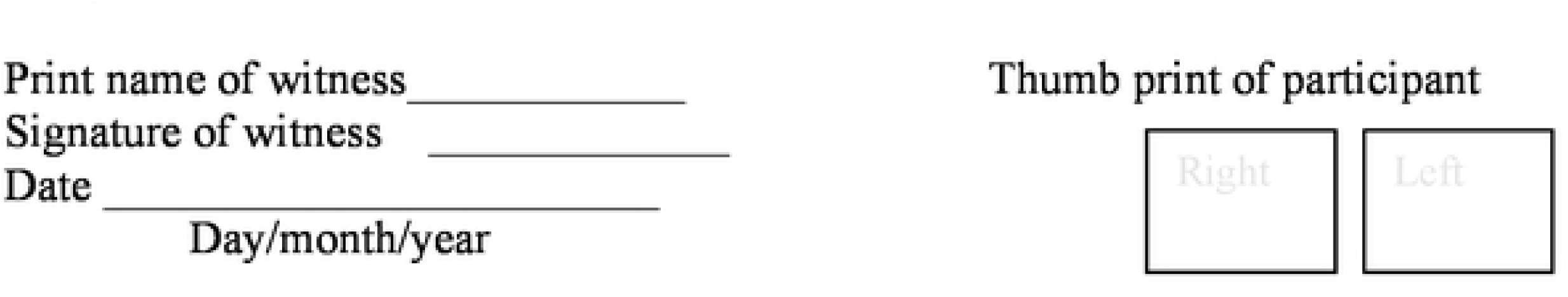

Statement by the rescarcher/pcrson taking consent

I have accurately read out the information sheet to the potential participant and to the best of my ability made sure that the participant understands

I confirm that the participant was given an opportunity to ask questions about the study, and all the questions asked by the participant have been answered correctly and to the best of my ability. I confirm that the individual has not been coerced into giving consent, and the consent has been given freely and voluntarily.

A copy of this ICF has been provided to the participant.

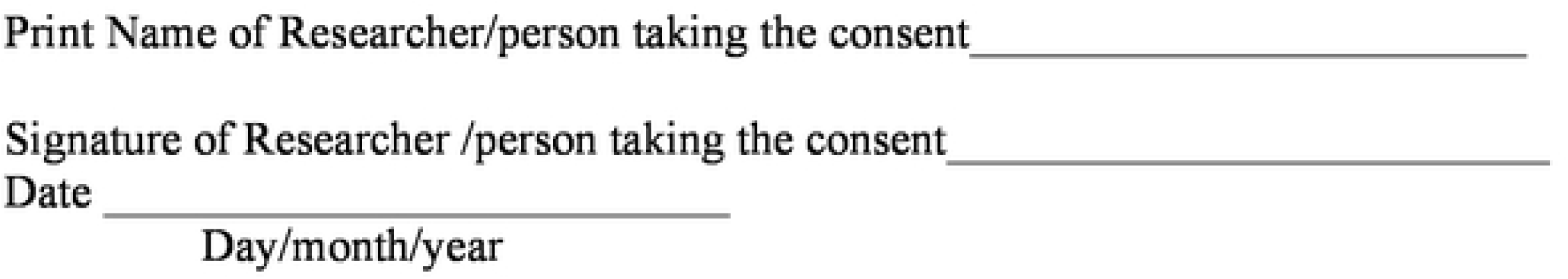

Phone Number: +8801749308699 (Saima Mehjabeen)

**Graph 3.**
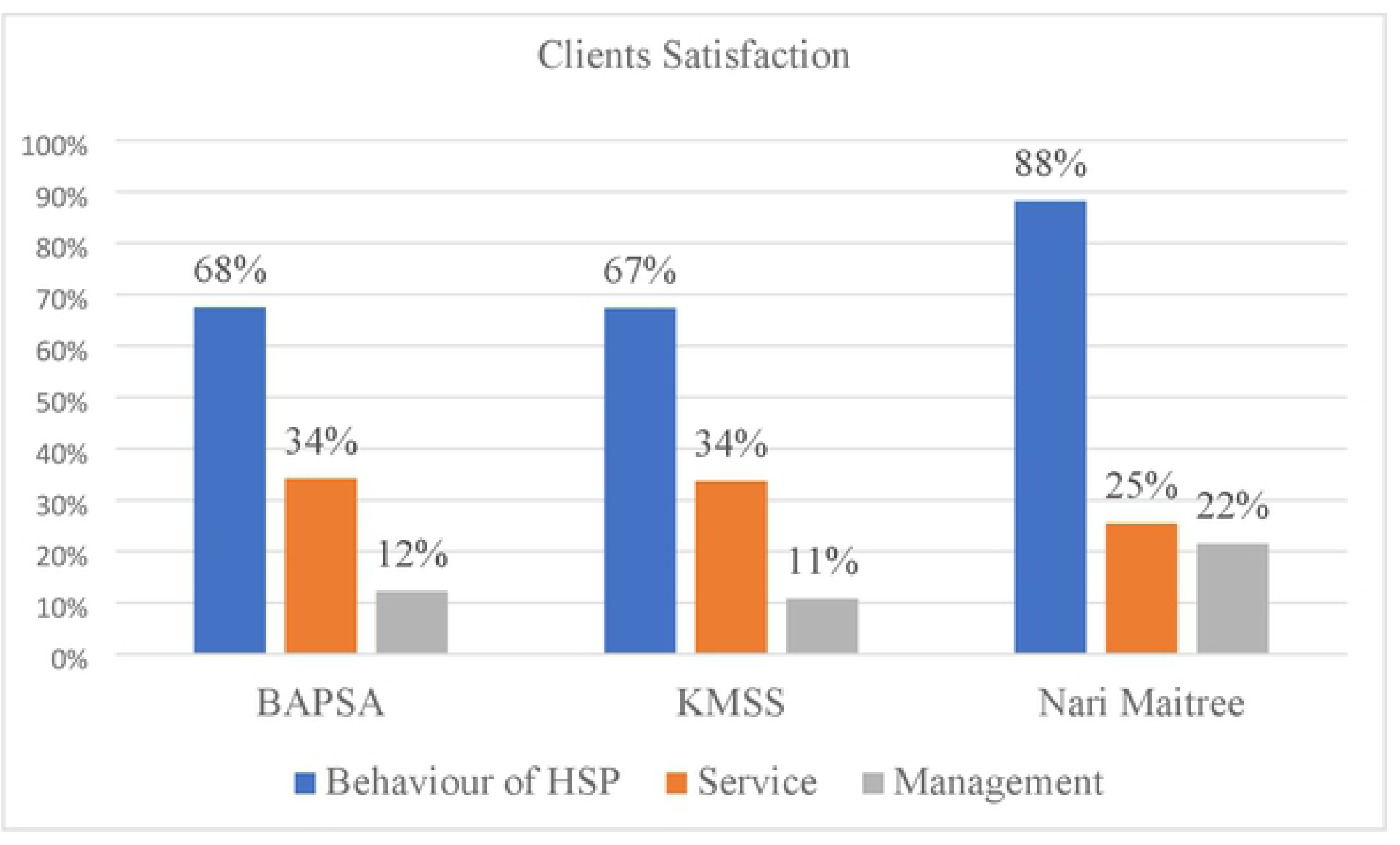
Clients satisfaction in various categories in three health facilities

**Graph 4.**
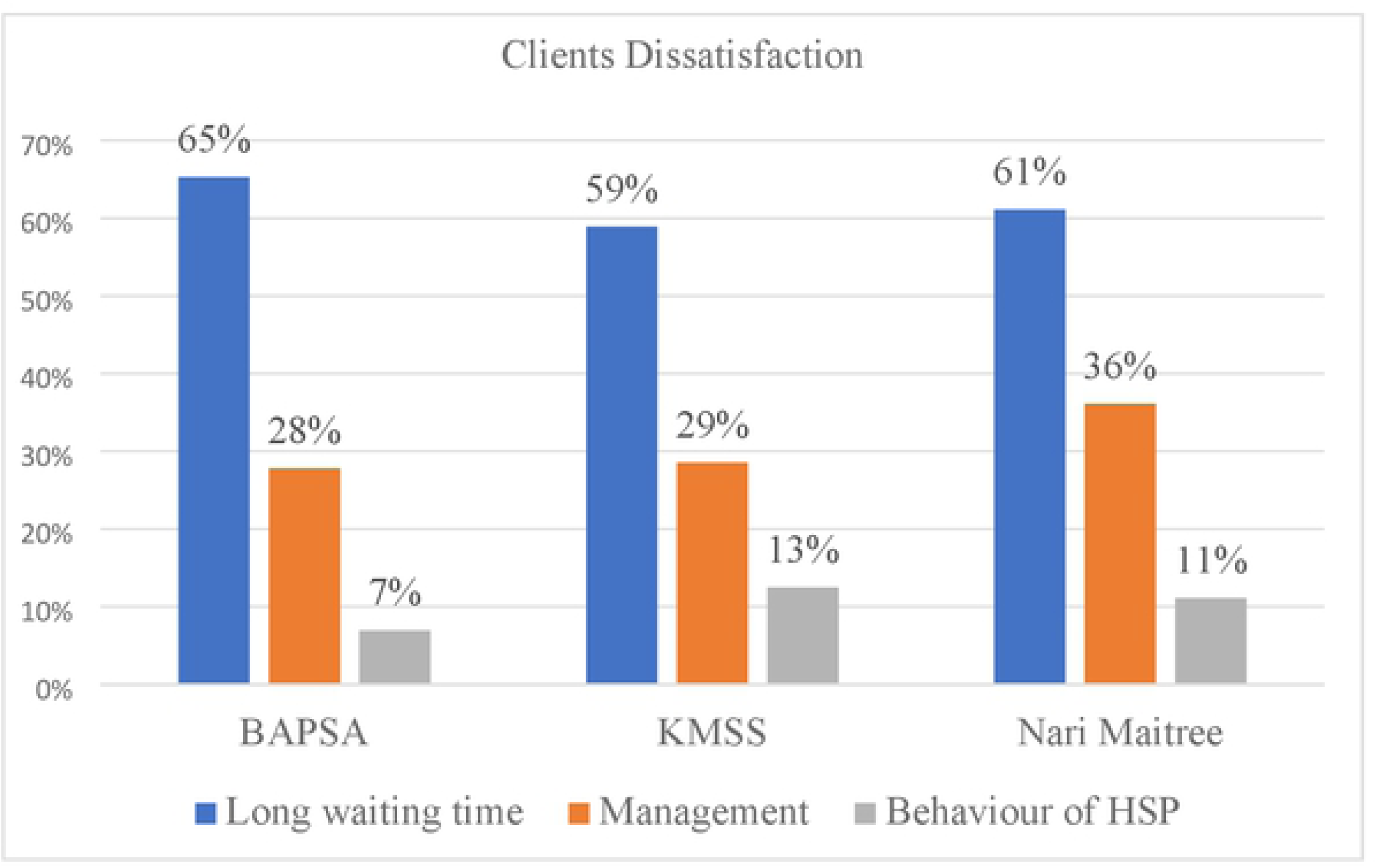
Client dissatisfaction about three health facilities

**Graph 5.**
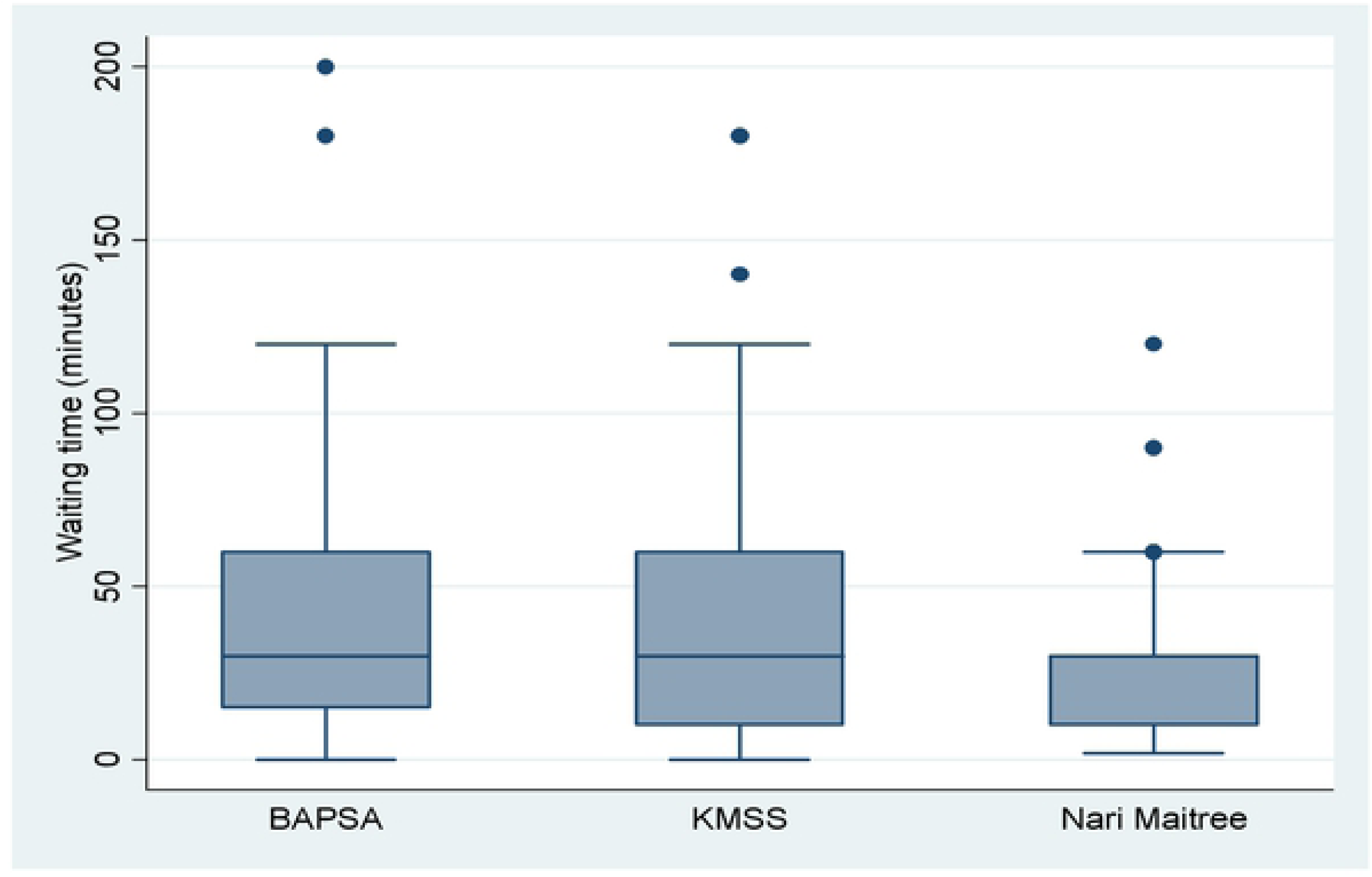
Box plot showing waiting time in three UPHCs.

## Notes

* A literate witness must sign (if possible, this person should be selected by the participant and should have no connection to the research team). Participants who are illiterate should include their thumb print as well.

